# Statistical Assessment of Biomarker Replicability using MAJAR Method

**DOI:** 10.1101/2022.12.08.22283210

**Authors:** Yuhan Xie, Song Zhai, Wei Jiang, Hongyu Zhao, Devan V. Mehrotra, Judong Shen

## Abstract

In the era of precision medicine, many biomarkers have been discovered to be associated with drug efficacy and safety responses, which can be used for patient stratification and drug response prediction. Due to small sample size and limited power of randomized clinical studies, meta-analysis is usually conducted to aggregate all available studies to maximize the power for identifying prognostic and predictive biomarkers. Since all available data are already aggregated, it is often challenging to find an independent study to replicate the discoveries from the meta-analysis (e.g., in meta-analysis of pharmacogenomics genome-wide association studies (PGx GWAS)), which seriously limits the potential impacts of the discovered biomarkers. To overcome this challenge, we develop a novel statistical framework, MAJAR (Meta-Analysis of Joint effect Associations for biomarker Replicability assessment), to jointly test prognostic and predictive effects and assess the replicability of identified biomarkers by implementing an enhanced Expectation–Maximization algorithm and calculating their posterior-probability-of-replicabilities (PPR) and Bayesian false discovery rates (Fdr). Extensive simulation studies were conducted to compare the performance of MAJAR and existing methods in terms of Fdr, power, and computational efficiency. The simulation results showed improved statistical power with well-controlled Fdr of MAJAR over existing methods and robustness to outliers under different data generation processes while considering both prognostic and predictive effects in the model. We further demonstrated the advantages of MAJAR over existing methods by applying MAJAR to the PGx GWAS summary statistics data from a large cardiovascular randomized clinical trial (IMPROVE-IT). Compared to testing main effects only, MAJAR identified 12 novel variants associated with the treatment-related LDL cholesterol (LDL-C) reduction from baseline.

## Introduction

In the era of precision medicine, many genetic biomarkers have been discovered to be associated with inter-individual drug response variability [1–4]. For instance, multiple genes have been shown to be associated with drug responses in terms of either efficacy or safety and are currently used in clinical practice, e.g., *APOE* with Statins therapy [5], *CYP2D6* with Tamoxifen treatment [6], and *HLA-B* with Abacavir hypersensitivity syndrome [7]. Due to small sample sizes from single randomized clinical studies, meta-analysis is usually conducted to aggregate all available studies to maximize the power of identifying novel prognostic and predictive biomarkers. Most analyses were conducted using traditional meta-analysis methods including fixed-effects meta-analysis [8], random effects meta-analysis [9], and their other adaptations [10,11]. However, since all available data are aggregated in meta-analysis for maximizing the power, these approaches often result in the lack of an independent dataset to replicate new discoveries (e.g., in pharmacogenomics genome-wide association studies (PGx GWAS)), which seriously limits the potential impacts of the discovered biomarkers and hampers the usage of these biomarkers in clinical practice. In addition, these traditional methods are usually sensitive to outliers in participating studies. To address these problems in disease genetic studies (i.e., disease GWAS), a model-based method MAMBA (Meta-Analysis Model-based Assessment of replicability) was recently proposed, which adopts a hierarchical mixture model to assess the posterior probabilities of replicability for identified variants by leveraging the strength and consistency of association signals among contributing studies [12]. However, this method is only designed to test the marginal prognostic (or genetic main) effects without considering predictive effects, thus it can be only applied in disease GWAS instead of PGx GWAS or other clinical settings. In disease genetic studies, the joint meta-analysis idea has been implemented for jointly testing genotype main effect and genotype by environment interaction effect and such joint meta-analysis methods have gained success in literature [13,14]. For example, Manning et al. propose a method to jointly test Single Nucleotide Polymorphism (SNP) and SNP-by-environment interaction effects, which is named JMA (Joint Meta-Analysis of SNP and SNP×E) [14]. Manning et al. then apply JMA to 60 SNPs within 1M base pairs (bp) of a gene (*PPARG*) that are found to be associated with type 2 diabetes. As BMI may modify the risk conferred by the variants in *PPARG*, they jointly test SNP and SNP-by-BMI interaction effects on a diabetes-related quantitative trait of fasting insulin levels in five cohorts. Their results show that meta-analyzing the main effect and interaction effect separately doesn’t identify any signals, while jointly testing both effects by using two degrees of freedom (2df) test from JMA identifies three signals. Although JMA considers both effects affecting the disease of interest, it is hard for it to quantify the replicability of identified variants like what MAMBA does. Therefore, it is of both theoretical interest and practical need to develop a MAMBA-like method to consider not only prognostic effects, but also predictive effects for the assessment of replicability in genetic biomarker analyses in randomized clinical trial settings (i.e., PGx GWAS).

In this paper, we propose MAJAR (**M**eta-**A**nalysis of **J**oint effect **A**ssociations for biomarker **R**eplicability assessment) to jointly test prognostic and predictive effects in meta-analysis without the need for using an independent cohort for replication of the detected biomarkers. MAJAR builds upon a two-dimensional hierarchical mixture model and adopts an efficiently-implemented Expectation-Maximization (EM) algorithm [15] for parameter estimation. It provides posterior probabilities of replicability (PPRs) and Bayesian false discovery rates (Fdr) for all biomarkers tested. Rather than relying on individual-level genotype and clinical data, MAJAR conducts meta-analysis across multiple studies based on summary statistics. Compared with methods directly using individual-level data across studies, methods using summary statistics better protect study participant privacy, reduce logistical difficulties and computational burden. Furthermore, MAJAR is computationally efficient due to the multiple speeding-enhancing strategies we implement in the MAJAR R package.

We conduct extensive simulations to compare the false discovery rate and power of MAJAR with existing methods across a wide range of genetic architectures and under different data generation processes. Our extensive simulation studies show that MAJAR outperforms other methods when both prognostic and predictive effects affect the phenotype of interest. We further demonstrate the utility of MAJAR by applying it to the PGx GWAS summary statistics data from a large cardiovascular randomized clinical trial (IMPROVE-IT) [16–18]. Under the threshold of Fdr < 0.05, we identified 13 variants associated with the treatment-related LDL cholesterol (LDL-C) reduction or change from baseline, 12 of which were missed by only testing for the main effect (i.e., while using MAMBA method). The top three identified variants with very high probabilities of replicability (PPR > 0.99) show biological evidence from multiple studies in the GWAS catalog [19]. These numerical studies demonstrate the advantage of MAJAR over the existing methods.

## Materials and methods

### MAJAR workflow

MAJAR is a statistical method proposed for jointly testing prognostic and predictive effects and assessing the replicability of identified biomarkers without using an independent cohort in the framework of meta-analysis. It can be used for analysis of general biomarkers such as clinical biomarkers, oncology biomarkers, genomics biomarkers, and other biomarkers. In this paper, we introduce MAJAR under the setting of analyzing genetic biomarkers in the context of PGx GWAS. The whole workflow of the proposed MAJAR method is summarized in Figure 1. The inputs of MAJAR are summary statistics of *M* SNPs from *K* studies. We denote the genotype (G) and genotype-by-treatment interaction (GT) effect size estimates as *b*_*jk,G*_ and *b*_*jk,GT*_ and the standard errors as *s*_*jk,G*_ and *s*_*jk,GT*_ for SNP *j* in study *k* (*k*=1, 2, …, *K*; *j*=1, 2, …, *M*). MAJAR takes the input summary statistics from *K* studies and uses an EM algorithm to estimate hyperparameters formulated in a two-dimensional hierarchical mixture model and calculates PPR for each SNP. For inference of replicable SNPs, MAJAR further converts the PPRs into Bayesian False discovery rate (Fdr) values using a joint local false discovery rate (Jlfdr) method previously proposed [20] to control the global Fdr under a specific threshold.

**Figure 1.**
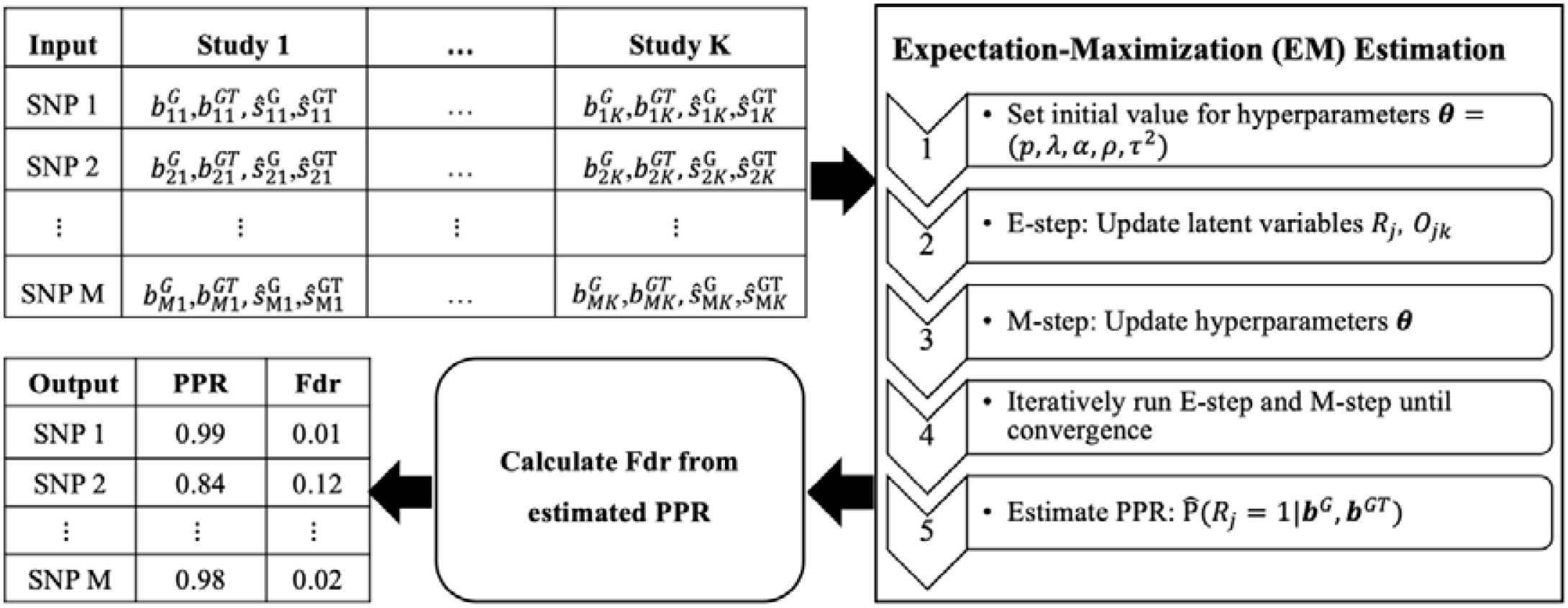
Overview of MAJAR method. Left top table: the summary statistics data from K studies including the genotype (G) and genotype-by-treatment (GT) effect size estimates and, and the standard errors as and for SNP *j* in study *k* (*k* = 1, 2, …, K; *j* = 1, 2, …, M). This summary statistics data is served as the input of the EM algorithm. Right panel: the EM algorithm procedure to estimate the parameters and posterior probability of replicability (PPR) for each SNP. Left bottom box: after the PPRs are estimated, the Fdr can be further calculated using the Jlfdr method. Left bottom table: the MAJAR algorithm outputs both the PPR and Fdr for each SNP.

### Two-dimensional hierarchical mixture model

MAJAR is a two-dimensional hierarchical mixture model extended from MAMBA [12] by considering biomarkers’ prognostic (G) and predictive (GT) effects simultaneously. In the hierarchical mixture model, there are two sets of latent variables *R*_*j*_ and *O*_*jk*_, where *R*_*j*_ is the indicator that SNP *j* has a real non-zero effect and *O*_*jk*_ is the indicator that a non-replicable zero effect SNP *j* is a spuriously associated outlier in study *k*. There are two assumptions for latent variables. 1) We assume replicable non-zero effect SNP *j* has the latent association status *R*_*j*_ in terms of the joint effect of G and GT effects, where *R*_*j*_ follows a Bernoulli distribution with the proportion of replicable non-zero effect SNPs being *p*. 2) We assume non-replicable zero effect SNP *j* in study *k* has the latent outlier status *O*_*jk*,_ where *O*_*jk*_ follows a Bernoulli distribution with the proportion of outlier zero-effect SNPs being *λ*. Given *R*_*j*_ and *O*_*jk*_, we assume that G and GT effect size estimates follow a mixture of bivariate normal distribution. When SNP *j* has a replicable non-zero effect, the mean of the bivariate normal distribution is 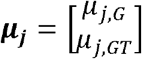. When SNP *j* is a non-replicable variant, the mean of the bivariate normal distribution is 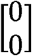. For spurious associations from non-replicable zero-effect variants, we further use an inflation factor *α* to characterize the extent of inflation in the observed effect sizes. To capture the correlation between G and GT, we assume the prior distribution of true effect sizes ***μ***_*j*_ follows a bivariate normal distribution, e.g., MVN(0, Ω), where 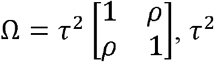 is the variance of true effect sizes of G and GT, and *ρ* is the correlation between true effect sizes of G and GT.

Together, we can summarize our MAJAR two-dimensional hierarchical mixture model as:

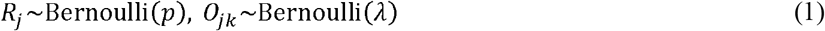

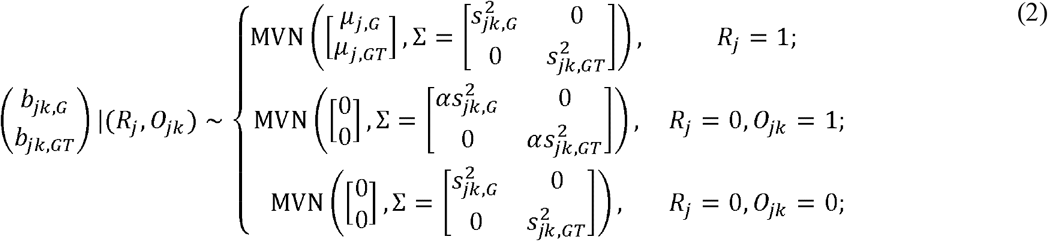

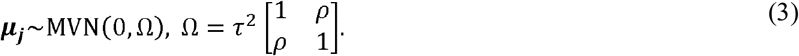

where *α* is an inflation factor that captures the extent of inflation in the observed effect sizes for outlier summary statistics and we denote the hyperparameters of MAJAR as ***θ*** = (*p, λ, α, τ*^2^, *ρ*).

### Estimation - EM algorithm

We develop an EM algorithm for hyperparameter estimation that is computationally efficient when handling variants on a genome-wide scale. To estimate the parameters, the EM algorithm runs Expectation (E-step) and Maximization (M-step) iteratively until convergence. At round *t*, it updates the latent variables 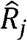 and 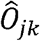 by their posterior probabilities given the parameter estimates at iteration *t* − 1, which is shown in equations 4 and 5.

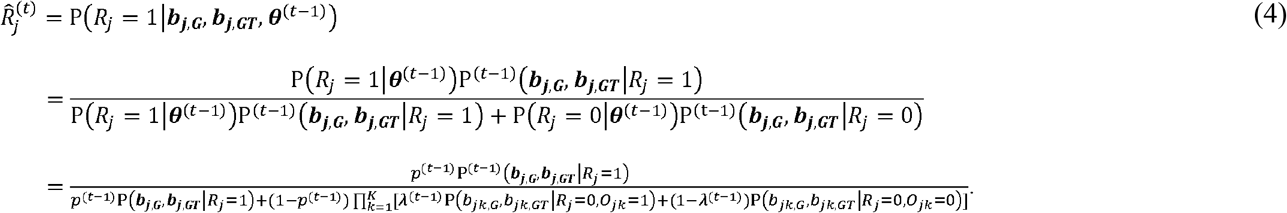

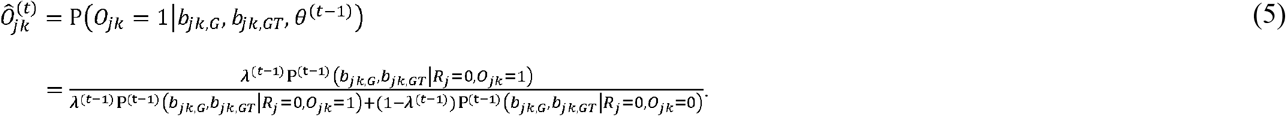

In the M-step, the hyperprior parameters ***θ*** are updated by maximizing the expected log-likelihood calculated from the E-step. For *p, λ* and *α*, there are explicit solutions, which are:

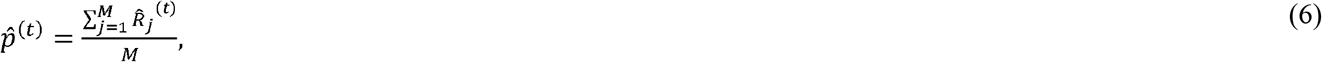

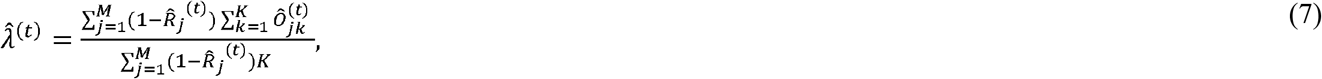

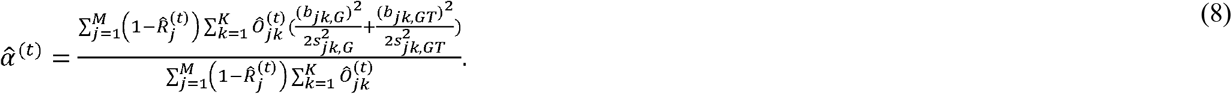

As there is no explicit solution for (*τ*^2^, *ρ)*, we utilize the unconstrained quasi-Newton method optimizer nlminb() [21] implemented in R [22] to optimize the two parameters by maximizing the terms related to them in the expected complete data log-likelihood (*ll*) (equation 9). More details of the derivations are described in the Supplementary Methods Section 1 (Optimizing (*τ*^2^, *ρ*) parameters).

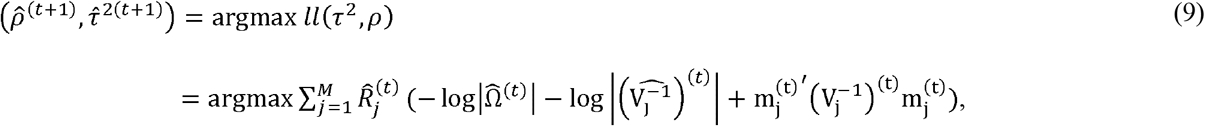

where 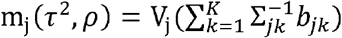 and 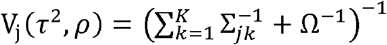.

After the EM algorithm converges and the hyperparameters are estimated, the PPRs of the tested SNPs can be estimated directly from the output of E-step as

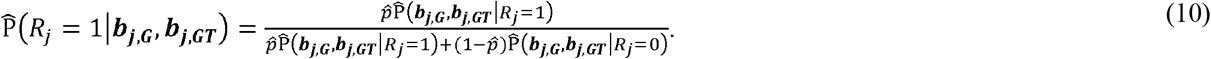

### Inference - Jlfdr approach

Jlfdr is defined as the posterior probability of a null hypothesis given the observed summary statistics. Here, the summary statistics are 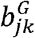 and 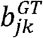 in the MAJAR model.

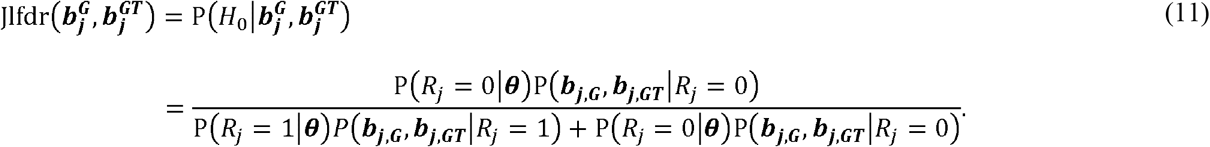

Jiang and Yu [20] have shown the relationship between Jlfdr and Fdr is

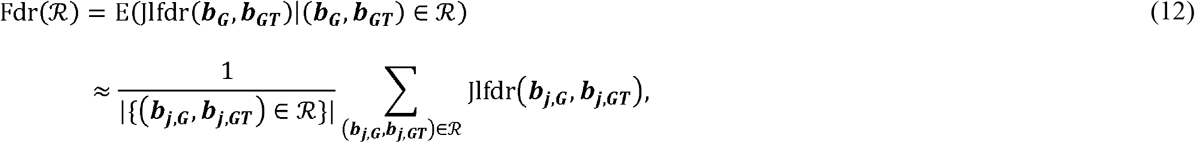

where the rejection region ℛ is the set of two-dimensional vectors ***b***_***j***,***G***_, ***b*** _***j***,***GT***_) that the null hypothesis can be rejected based on specific rejection criteria. Based on the derivation in the original method [20], the most powerful rejection region for a given Fdr level q is {Jlfdr (***b***_***j***,***G***_, ***b*** _***j***,***GT***_ ≤ *t* (*q*)}, where *t* (*q*) is a threshold that can be determined by Fdr(ℛ^*a*^) = *q*. The threshold of *t*(*q*) can be determined by first sorting the Jlfdr values of all tested SNPs in ascending order with the a-th Jlfdr value as Jlfdr^a^, and then approximating the Fdr of the region as

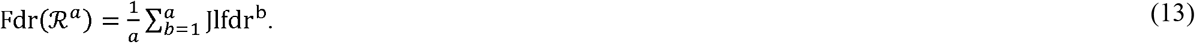

Denote c=max{a| Fdr(ℛ^*a*^) ≤ *q*)}, then the Jlfdr threshold *t*(*q*) is Jldfr^c^. The decision rule is to reject all SNPs with Jlfdr 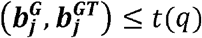.

### Fast Implementation of the MAJAR Algorithm

Motivated by MAMBA [12], we have implemented MAJAR with three strategies to increase its computational efficiency: 1) using the mcapply() function in the R parallel package for multi-core computation to update *O*_*jk*_ by running *M* SNPs in parallel; 2) implementation of E-step functions using C++ through the R Rcpp package [23,24] and M-step optimization function (equation 9) with the R TMB package [24]; and 3) unbounding the parameter space of (*τ*^2^, *ρ*) when maximizing the expected log-likelihood. Specifically for 2) the MakeADFun function in the package can calculate first and second order derivatives of the likelihood function written in C++ and the function and its derivative can be called from R. For 3), we map *ρ* to arctanh (*ρ*) to unbound the parameter space from (−1, 1) to (−Inf, Inf), and map *τ*^2^ to log *τ*^2^ to unbound the parameter space from (0, Inf) to (−Inf, Inf). We conducted a benchmark analysis to compare the computation time of our method with and without implementing these strategies. The fast implementation of our software can be accessed from our GitHub webpage https://github.com/JustinaXie/MAJAR.

### Simulation studies

We conducted extensive simulation studies to benchmark the performance of MAJAR with other commonly used meta-analysis and replicability analysis methods including meta-analysis fixed effect model (FE), meta-analysis random effect model (RE), and MAMBA. We first checked whether MAJAR can estimate the parameters accurately under the ground truth model and then compared the power performance of MAJAR with MAMBA, FE, and RE under the data generation process of MAJAR, FE and RE’s ground truth models, respectively. Given FE, RE, and MAMBA can only perform marginal analysis instead of conditional or joint effect analysis, we inferred replicable SNPs by testing only the marginal G effect for these three methods. We also verified whether MAJAR simplifies into MAMBA when only the G effect exists.

We randomly simulated 5,000 SNPs in total and varied the number of studies *K* in the meta-analysis from 2, 5, to 10. We used standard errors estimated from the GWAS summary statistics of our real (IMPROVE-IT) PGx data. The proportion of replicable non-zero effect SNPs *p* was set to 0.05, 0.1 or 0.2. According to the relationship between the variance of SNPs (*τ*^2^) and the heritability 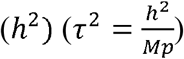 [25], we varied *τ*^2^ by setting *h*^2^ from 0.1, 0.3 to 0.5.

Under the data generation process of the MAJAR ground truth model, we set the proportion of non-replicable zero-effect SNPs *λ* to 0.1 and varied the inflation factor *α* from 1, 2, 5, to 15. We also tested the robustness of MAJAR under different correlation settings of G and GT by varying the correlation parameter *p* from -0.75, -0.25, 0, 0.25, to 0.75. The simulation procedure under MAJAR’s ground truth model includes three steps. In the first step, we simulated the latent variables *R*_*j*_ ∼ Bernoulli (*p*) and *O*_*jk*_ ∼ Bernoulli (*λ*). In the second step, we simulated the true effect sizes 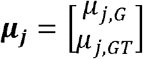 with a spike-and-slab prior. That is, when *R*_*j*_ = 0, ***μ***_***j***_ **= 0;** and when *R*_*j*_ = 1,

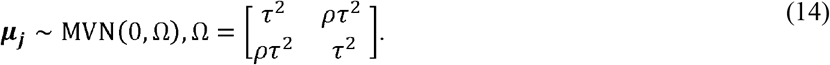

In the third step, we simulated the effect size estimates 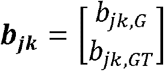 from the mixture bivariate normal distribution according to equation 2. Simulation details under the FE and RE data generation process are introduced in Supplementary Methods Section 2-3. Robustness analysis was also conducted under an extended model of MAJAR in which the variance *τ*^2^ of G and GT are not the same (i.e., 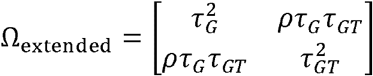).

We replicated the simulation 100 times under each setting. The false discovery proportion (FDP) was defined as the proportion of false rejections and the expected value of FDP given the number of rejections is larger than 0. We set the Fdr threshold as 0.05 for discovery of replicable SNPs. Computation time was benchmarked using 2.4 GHz Intel Core i5 with 4 parallel cores for all methods compared.

### Application to IMPROVE-IT PGx GWAS data

We applied MAJAR to jointly testing the prognostic and predictive effects on the low-density lipoprotein cholesterol (LDL-C) log-fold change at 1 month in a PGx GWAS summary statistics data. This data was from the IMPROVE-IT (IMProved Reduction of Outcomes: Vytroin Efficacy International Trial, clinical trial registry number: NCT00202878) study. IMPROVE-IT is a multi-center, double-blind, phase 3b randomized trial to establish the efficacy and safety of Vytorin (ezetimibe□+□simvastatin tablet) versus simvastatin monotherapy in high-risk subjects [16]. A detailed introduction of the study population, genotyping, genotype quality control (QC), and imputation for the GWAS analyses were described in previous studies [17,18]. After variant-level QC and imputation, there were 9,407,967 variants and 6,502 subjects available for analysis. After filtering out subjects who had a cardiovascular event prior to our analysis time point (month 1) to avoid interference unrelated to treatment on LDL-C, a total of 5,661 unrelated European subjects were included in the final analysis. Similar to the simulations, we also applied MAMBA to testing the prognostic (G) effect and predictive (GT) effect separately in the analysis of the IMPROVE-IT PGx GWAS summary statistics data.

Due to the lack of PGx GWAS data from multiple studies with the same endpoint, we proposed an alternative strategy to analyze the IMPROVE-IT data by randomly splitting the whole cohort into five folds. We conducted GWAS on each fold and used the summary statistics from each fold, only keeping variants with MAF > 0.01 as input for MAJAR. To boost the computational speed, we adopted a similar strategy as MAMBA [12] to clump variants with suggestive evidence from a reference method, prune variants with relatively low evidence, and then combine variants together in the two sets. Cauchy combination test (CCT) [26] was used to get combined *p* -values, which were further used as a reference for clumping and pruning in PLINK 1.90. For clumping, we used parameters “–clump-p1 1e-4 –clump-kb 1000 –clump-r2 0.1”. For pruning, we used 1000 genome European samples [27] as a reference panel with parameters “–indep-pairwise 1000 kb 1 0.1” and matched the pruned sets with the IMPROVE-IT variants. To ensure the combined sets of variants are not within the same linkage equilibrium (LD) block, we removed all pruned variants in 1M bp within clumped variants. Then, we ran MAJAR, MAMBA, FE, and RE on the variants left for each chromosome to allow different parameters estimated for each chromosome. In practice, we set the inflation factor *α* ≥1 for both MAJAR and MAMBA and set the initial hyperparameter values of MAMBA (*p, λ, α τ*^2^) the same as those of MAJAR to make a fair comparison. We followed the same penalty strategy from previous studies [20,28]. We also conducted the interaction test of the GT effect with MAMBA by using *b*_*jk*_ and *s*_*jk,GT*_ as input without conditioning on the G effect. We further pooled PPRs from all chromosomes, converted them to Fdr values, and compared the results with CCT *p*-values and the pooled GWAS 2df *p*-values [17]. In addition, to examine the robustness of the results, we also conducted a sensitivity analysis using a five-fold cross-validation procedure, for example, by leaving one fold out from the five folds and repeating the same analytical procedures on the remaining four folds.

## Results

### Simulation results: Fdr and power under ground truth model

Before evaluating the Fdr and power under the ground truth model (i.e., data generation process or DGP = MAJAR), we first checked whether MAJAR can accurately estimate the true effect sizes (*μ*_*G*_, *μ*_*GT*_) and hyperparameters (*p, λ, α, ρ, τ*^2^). The simulation results are summarized in Figure S1, which shows that MAJAR has generally unbiased estimates under different combinations of the number of studies and the heritability. It is worth noting that the mean square error (MSE) of *μ*_*G*_ and *μ*_*GT*_ slightly increased when the heritability increased. This is as expected since the magnitude of genetic effect sizes was larger when the genotype explained more variances in the phenotype. Overall, the MSEs were very small (i.e., close to 0 at 10^−3^ level), indicating that MAJAR can accurately estimate the true effect sizes and hyperparameters.

Given the unbiased estimation, we further visualized the Fdr and power results for comparing FE, RE, MAMBA, and MAJAR under different combinations of the number of studies *K*, the proportion of replicable non-zero effect SNPs *p*, and the heritability *h*^2^ by fixing inflation factor *α* at 5 and *ρ* at 0.75 (Figure 2). The simulation results in Figure 2 showed that FE was not able to control Fdr at the desired threshold of 0.05 under all settings, and RE also had slight inflation when both the study number *K* and the heritability *h*^2^ were small. In contrast, both MAMBA and MAJAR controlled Fdr well.

**Figure 2.**
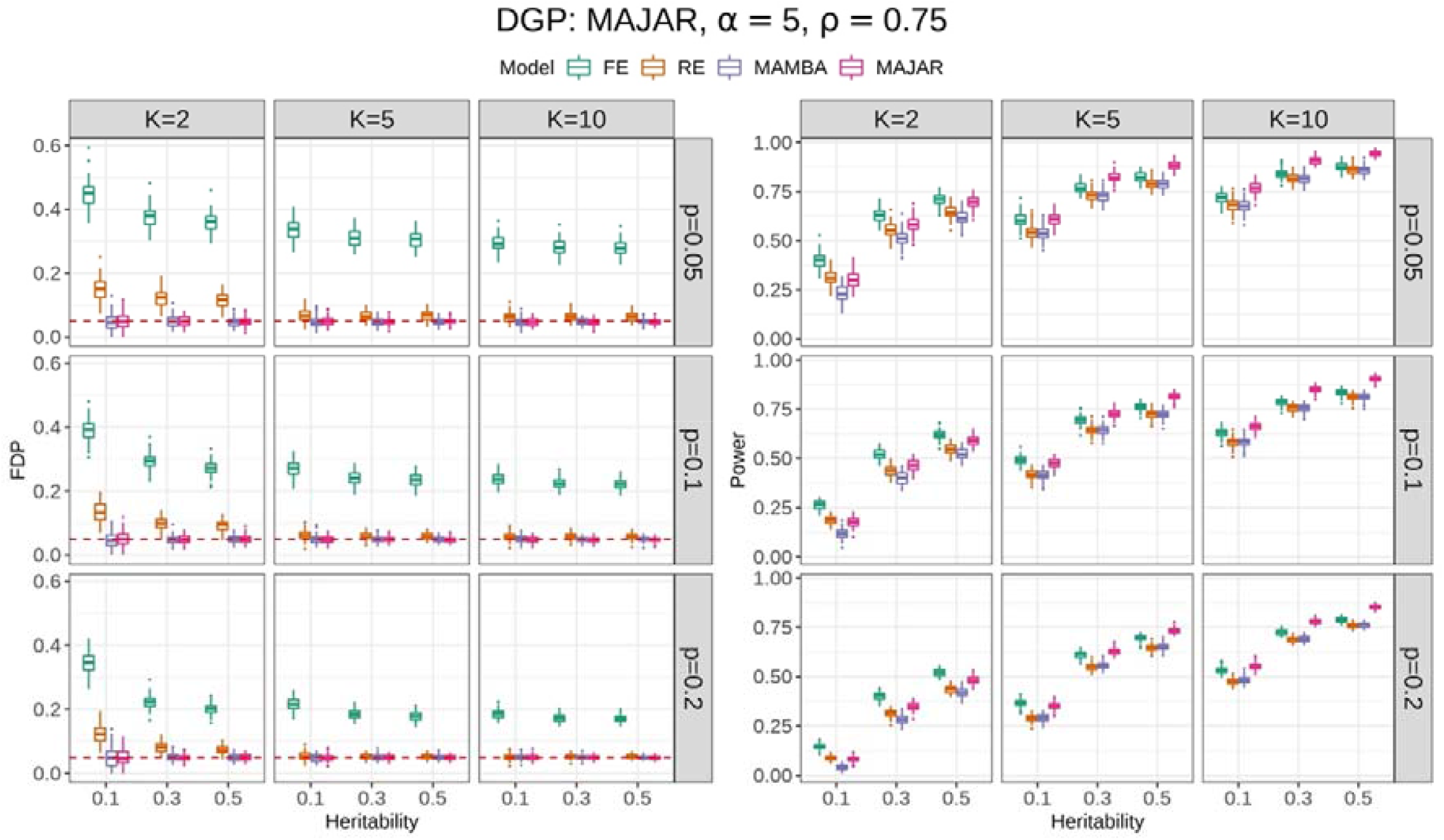
Simulation results of FDP and power while comparing FE, RE, MAMBA, and MAJAR under MAJAR’s ground truth model. The left panel is for the FDP (false discovery proportion) performance, and the right panel is for the power performance. For each panel, the columns represent the number of studies *K*, and the rows represent the proportion of replicable non-zero effect SNPs *p*. Each panel shows the FDP and power (y-axis) with increasing heritability *h*^2^ (x-axis) under different settings of *K* and *p*. The performance of each method in 100 replications under each simulation setting is visualized by the colored boxplots. The red dashed line in the FDP panel indicates the target Fdr threshold of 0.05.

For power comparisons, FE was not considered for the method comparison given its severe Fdr inflation (Figure 2, left panel). Overall, RE had comparable power to MAMBA, while MAJAR consistently had the best power among all circumstances (Figure 2, right panel). The power increased for all models as the study number *K* increased. However, the power improvement of MAJAR over other methods was even greater. For example, when *K* = 2, the power increase from RE to MAJAR was between 2.6% and 10.0%; when *K* = 10, such power increase was between 9.8% and 16.2% in terms of the median values under different combinations of *p* and *h*^2^. This indicated that MAJAR was more effective in leveraging information across multiple studies than other methods by modeling both prognostic and predictive effects. With the increase of the proportion of replicable non-zero effect SNPs *p*, the power decreased for all models. This is because the signal of non-zero effect SNPs became weaker given the relationship between the variance of true effect sizes 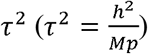. In that condition, we observed that the power of MAJAR decreased much slower than RE and MAMBA (e.g., MAJAR outperformed RE by increasing the power between 2.6% and 11.7% when *p* = 0.05 and between 8.5% and 16.2% when *p* = 0.2 in terms of the median values under different combinations of *K* and *h*^2^), suggesting that MAJAR is more robust to highly polygenic genetic architectures with small signal-to-noise ratio. Lastly, when the heritability increased, MAJAR was also superior to other methods (e.g., MAJAR outperformed RE by increasing the power between 2.6% and 13.1% when *h*^2^ = 0.1 and between 6.1% and 16.2% when *h*^2^ = 0.5 in terms of the median values under different combinations of *K* and *p*).

### Simulation results: Fdr and power under other DGPs

In the previous section, we showed that MAJAR had its advantages over other methods under the ground truth model (i.e., when DGP = MAJAR). In this section, we explore whether MAJAR still outperforms the existing methods under other DGPs. Under the data generation processes of FE and RE, we visualized the simulation comparison results of Fdr and power under different settings *K, p* and *h*^2^ and fixed *ρ* at 0.75 (Figure S2-S3). For data generated under FE, all methods controlled Fdr well except that the RE method tended to be a little conservative. The power performance of MAMBA was comparable to the FE method and MAJAR performed the best. For data generated under RE, the FE method had severe Fdr inflation, while other methods generally controlled Fdr well. MAMBA yielded better or comparable performance to RE, while MAJAR provided better performance than MAMBA. In a nutshell, MAMBA and MAJAR were more robust than FE and RE in terms of Fdr control under different DGPs, with MAJAR having superior power across different scenarios. Under the data generation process of MAMBA with prognostic effect only, we observed that MAJAR had a very similar false discovery proportion (FDP) and power performance as MAMBA (Figure S4). Overall, the power of MAJAR was slightly smaller than MAMBA when the number of studies was small. This is as expected since MAJAR, using a two-dimensional hierarchical mixture model to estimate prognostic and predictive effects simultaneously, would inevitably include noises when the predictive effect didn’t exist. When there were only G effects (i.e., without any GT interaction effects) and the number of studies was large, MAJAR had very similar power as MAMBA.

### Simulation results: sensitivity analyses

To test the robustness of MAJAR with regards to different correlations between G and GT effects *ρ*, we ran the simulations by following the MAJAR data generation process and fixing *α* at 5 and *h*^2^ at 0.3 and compared the power performance with increasing values of *ρ* (Figure S5). The results showed that the power of MAJAR first increased and then decreased with the increase of *ρ* and MAJAR had slightly better power performance when the correlation was weak regardless of the correlation directions (i.e., with a small absolute correlation). Overall, MAJAR was robust to the varied correlations between G and GT effect sizes.

To test the robustness of MAJAR with regards to different outlier inflation factor *α*, we fixed *ρ* at 0.75 and compared the power performance under different *α* (Figure S6). MAJAR controlled Fdr well under all settings. The power of MAJAR decreased with increasing *α* when the number of studies was small. This indicated that it was hard for MAJAR to differentiate the outliers among studies when there were only a small number of studies. When the number of studies was larger, MAJAR’s power under different *α* became comparable.

Under the MAJAR ground truth model, we assumed the prognostic (G) and predictive (GT) effect sizes were on the same scale by setting the variances of the two effects to the same *τ*^2^ value as in our previous simulation analyses. We also tested the robustness of our method under an extended model of MAJAR where variances of G and GT were different. In this simulation, we fixed the value of the heritability of 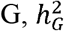, so the increase in the ratio 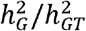 means a decrease in the heritability of 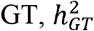. The simulation results are summarized in Figure S7. In summary, the simulation results showed that MAJAR had slight Fdr inflation and slightly decreased power when *K* = 2 and 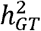 was much smaller than 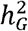 (i.e., the assumption was violated). With the increase of *K*, MAJAR had improved power with well-controlled Fdr, regardless of whether the assumption was violated or not.

### Application to IMPROVE-IT PGx GWAS data

After applying MAJAR to IMPROVE-IT PGx GWAS summary statistics data using our proposed five-fold strategy, we summarized the whole genome SNPs’ PPR and their corresponding Fdr in a scatter plot (Figure 3a) and further visualized the Fdr results in a Manhattan plot (Figure 3b). We determined the Fdr cutoff that corresponded to PPR > 0.99 (Figure 3a) and drew the corresponding blue line (Fdr < 0.003) on the Manhattan plot. In total, we observed 3 variants (rs1065853, rs56746789, rs62531463) under a stringent threshold of PPR > 0.99 (i.e., Fdr < 0.003) (Table 1). The most significantly associated variant (rs1065853) was located in *APOE/APOC1*, which is a well-known locus that is associated with response to statins and involved in lipid metabolism and drug transport [29,30]. This variant had very high MAMBA-G (MAMBA using summary statistics of G effect only as input) PPR (PPR=1) and was also a strong signal in the pooled GWAS 2df test (*p* = 1.09 × 10^−51^). The other two variants with PPR > 0.99 had relatively high PPRs for both MAMBA-G effects (rs56746789 PPR = 0.832; rs62531463 PPR = 0.638) and MAMBA-GT effect (rs56746789 PPR = 0.510; rs62531463 PPR = 0.439), indicating the boosted power of replicability from the GT effect added. The two variants are located in locus *LINC01482* and gene *KANK1*, respectively. By searching the *LINC01482* locus in the GWAS catalog (https://www.ebi.ac.uk/gwas/), we found there were in total 53 studies related to this locus and 33 associations between SNPs within this locus and LDL, HDL, and other lipid-related traits. By searching the *KANK1* locus on the GWAS catalog, we found there were in total 33 studies related to this locus and 2 associations related to lipid measurement. Previous studies have also reported the loci in *KANK2, KANK1*’s close paralog, are associated with LDL-C [31,32].

**Table 1.**
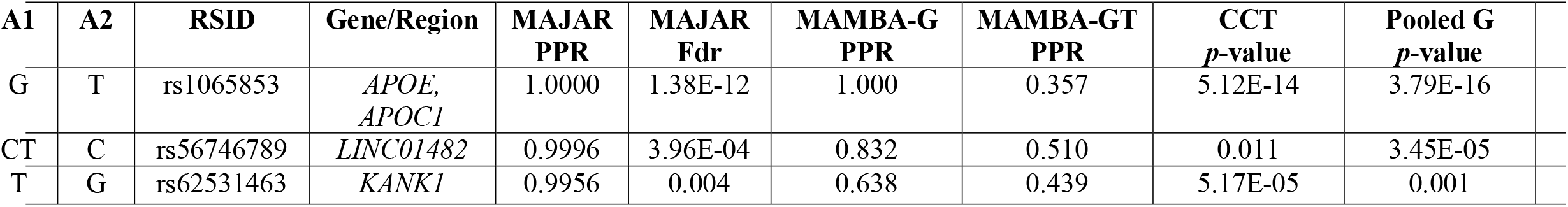
Top variants identified by MAJAR with PPR > 0.99 in the analysis of IMPROVE-IT PGx GWAS data and their results and annotation information.

**Figure 3.**
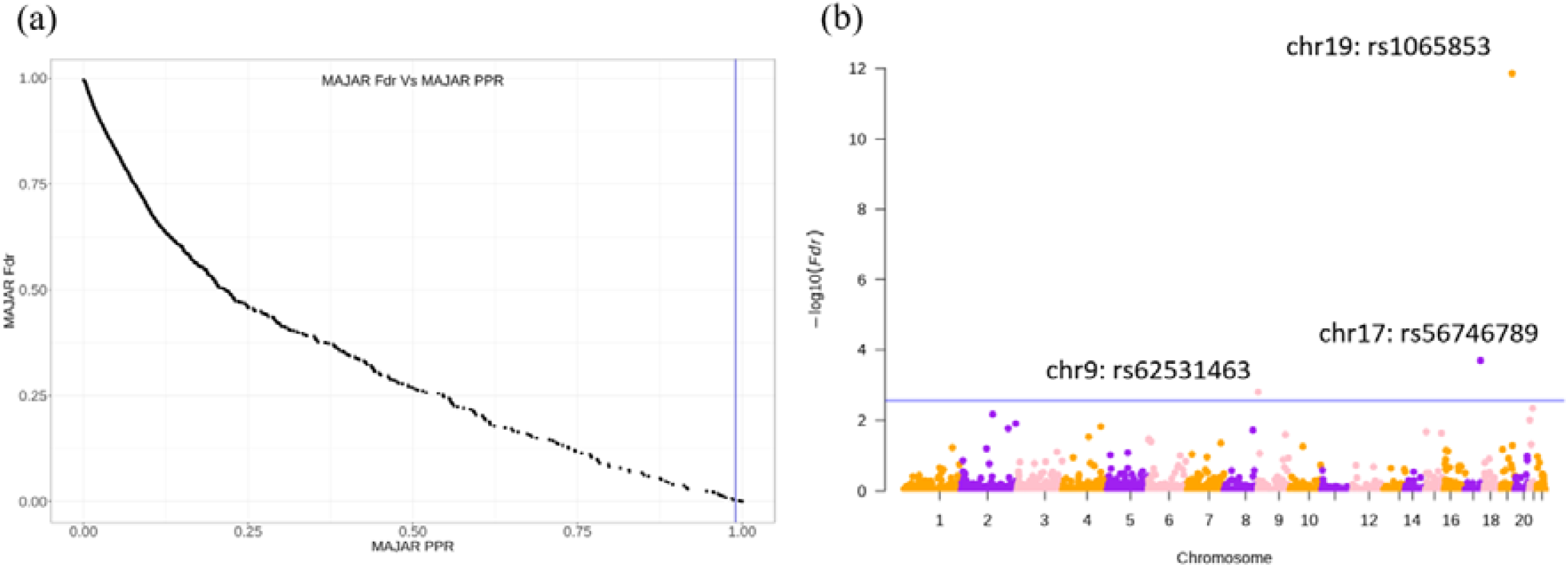
MAJAR whole genome meta-analysis results from the analysis of IMPROVE-IT GWAS summary statistics data. (a) Relationship between MAJAR Fdr and MAJAR PPR. The x-axis represents MAJAR PPR, and the y-axis represents MAJAR Fdr. The vertical blue line represents MAJAR PPR=0.99. (b) Manhattan plot for all tested variants. The y-axis represents the values, and the x-axis shows the genome position of all variants. The blue horizontal line represents the PPR threshold of 0.99 (Fdr = 0.003) determined from the relationship between Fdr and PPR shown in the left figure.

We further tested the robustness of the results in the framework of five-fold cross-validation after leaving one fold out from the five folds each time. We visualized the robustness analysis results based on the 5-fold cross-validation procedure in an upset plot (Figure S8). From the bottom right part of the upset plot, all three identified variants were consistently selected in at least 4 out of the 5 iterations. This demonstrated the robustness of the top 3 variants identified by MAJAR in the IMPROVE-IT data analysis. We identified 10 additional variants under the threshold of Fdr < 0.05 (Table S1) including two variants on chromosome 21 (rs76451912, rs282368), three variants on chromosome 2 (rs4954144, 235349598-C-T, rs35421171), two variants on chromosome 15 (rs28510330, rs201822759), and one each on chromosome 4 (rs1351220), chromosome 8 (rs35903937), and chromosome 9 (rs61381472). rs76451912 is located in the noncoding region. The nearest coding gene is *ADAMTS5*, which has been reported to have multiple associated SNPs with blood protein measurement in two studies [33,34] and a disintegrin and metalloproteinase with thrombospondin motifs 5 measurement, the proteolytic activity of which has been shown to reduce the LDL binding ability of biglycan and releases LDLs from human aortic lesions [35], in three studies [36–38]. rs4954144 is an intron variant in *MGAT5*, which has been reported to have multiple associated SNPs with multiple cholesterol related measurements [39]. The remaining top hits identified may represent novel genetic associations, which need further interpretation and exploration. The evidence of mapped genes of top variants downloaded from the GWAS catalog is shown in Table S2.

We also compared the PPR distributions among MAJAR, MAMBA-G, and MAMBA using summary statistics of GT effect only as input (MAMBA-GT). All three PPRs showed right-skewed distributions with long right tails, indicating that most SNPs were not replicable (non-zero effect) signals. The corresponding frequency dropped more smoothly when PPR increased for MAJAR method, compared to MAMBA-G and MAMBA-GT (Figure S9). We visualized the overlapping results among MAJAR, MAMBA-G, MAMBA-GT, and the 2df test from the pooled GWAS in terms of signal detection using Venn diagrams (Figure S10). Under the condition that the pooled GWAS 2df test *p*-values < 1 × 10^3^, MAJAR identified three overlapping variants (PPR > 0.99) with the pooled GWAS results, out of which one signal is also significant in the MAMBA-G analysis (i.e., PPR > 0.99). MAMBA-GT did not identify any variants with PPR > 0.99. After we used a more stringent threshold of the 2df test *p*-value < 1 × 10^−4^ for the pooled GWAS analysis, MAJAR identified two out of three previously identified overlapping variants with the pooled GWAS. Moreover, we checked the pair-wise consistencies between MAJAR PPRs and MAMBA-G PPRs, MAMBA-GT PPRs, − log_l0_ p-values from the pooled GWAS and − log_l0_ p -values from the CCT test, respectively. From the scatter plots, we observed clearly positive correlations between the PPR results from MAJAR and the results from existing methods (Figure 4). This also indicates the consistency trend of the signals discovered by different methods.

**Figure 4.**
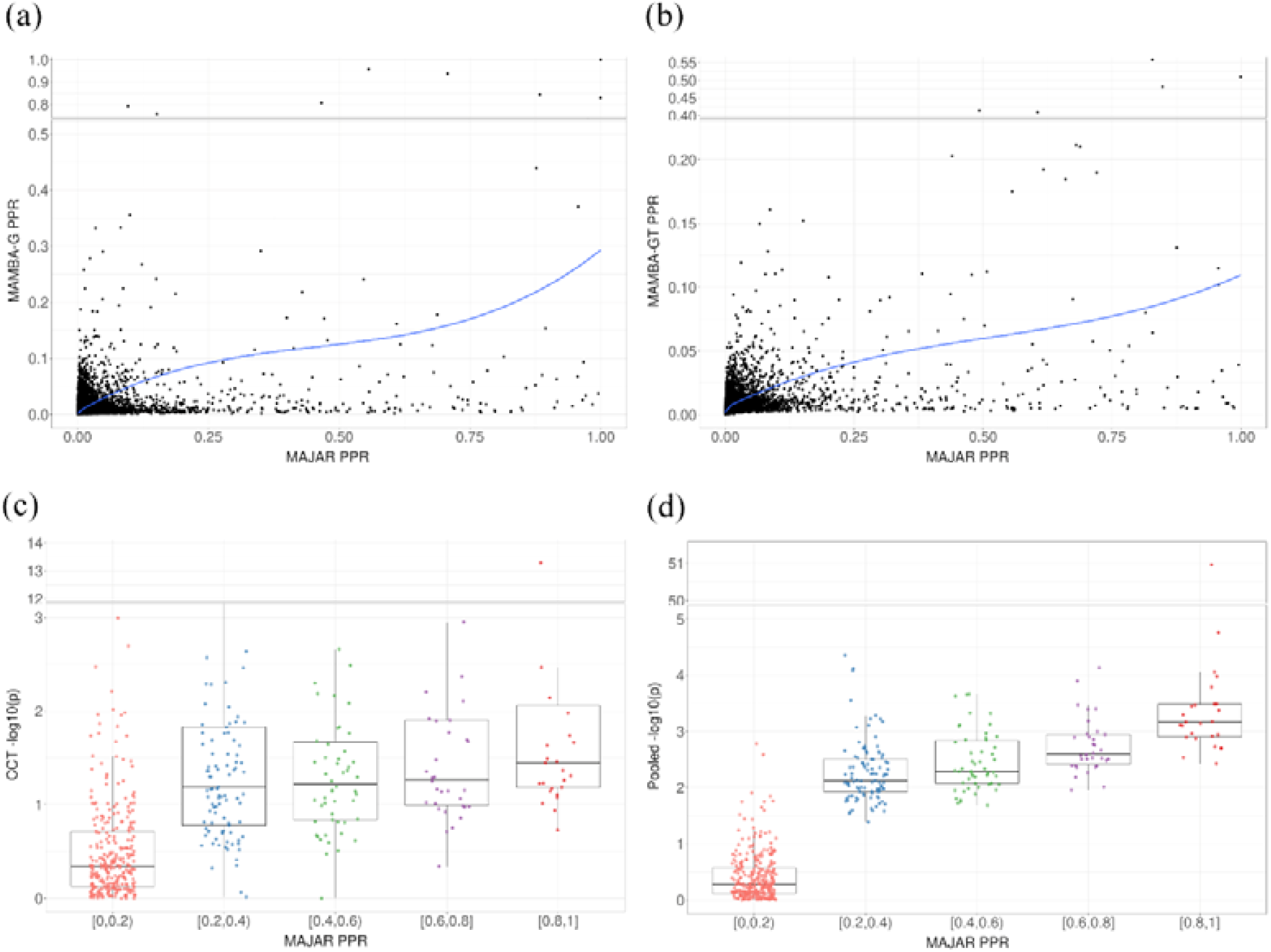
Comparison of MAMBA-G PPR, MAMBA-GT PPR, CCT *p*-values, pooled GWAS 2df test *p*-values versus MAJAR PPR. (a) Scatter plot of MAMBA-G PPRs versus MAJAR PPRs. The blue curve is fitted by the LOWESS smoother. (b) Scatter plot of MAMBA-GT PPRs versus MAJAR PPRs. The blue curve is fitted by the LOWESS smoother. (c) Scatter plot of CCT *p*-values and MAJAR PPRs. Variants are categorized and colored by five different bins of MAJAR PPRs. The boxplots demonstrate the distribution of CCT *p*-values under five different bins of MAJAR PPRs. (d) Scatter plot of pooled GWAS 2df *p*-values and MAJAR PPRs. Variants are categorized and colored by five different bins of MAJAR PPRs. The boxplots demonstrate the distribution of pooled GWAS 2df *p*-values under five different bins of MAJAR PPRs.

### Computation time

We compared the computation time of MAJAR with FE, RE, and MAMBA under the MAJAR data generation process (Materials and Methods). Empirically, we observed the computational bottleneck of MAJAR’s algorithm is the estimation of (*τ*^2^, *ρ*) in the M-step. When only R code is used to implement the algorithm, the computation time is much slower than the efficient implementation with parallel computing and C++. The computation time comparison results were summarized in Figure S11. From this figure, we observed that the FE model was the fastest under all simulation settings followed by RE, which is due to its model simplicity. MAMBA was slower than FE and RE. This might be due to the relatively more stringent convergence criteria of the algorithm. The fast version of MAJAR (partly written in C++ and implemented with 4 cores in parallel) was comparable to RE with the increase of *K*. Theoretically speaking, the proposed MAJAR model (i.e., two-dimensional hierarchical mixture model) was more complicated than MAMBA since it has more hyperparameters to be estimated in the EM algorithm. Figure S11 shows that MAJAR runs much faster than MAMBA, which indicates that our fast implementation strategies can indeed improve the computational efficiency. In fact, MAJAR without fast implementation (written in R and implemented with only one core) was much slower than the other methods: MAJAR without fast implementation was around seven times slower than MAMBA and 20 times slower than MAJAR with fast implementation. Under most simulation settings, the fast version of MAJAR could perform the test for 5,000 SNPs within a minute.

For the IMPROVE-IT real data application, we conducted the analysis by parallelizing the 22 chromosomes, so that the time cost is the computation time of the largest chromosome. In our data, the chromosome that required the longest time was chromosome 2 with 11,213 SNPs after pruning and clumping. The running time of MAJAR was 9.3 minutes, MAMBA-G 34.7 minutes, and MAMBA-GT 35.4 minutes. These results demonstrate that MAJAR can efficiently conduct the assessment of biomarker replicability in studies with a large number of biomarkers such as in PGx GWAS.

## Discussion

In this study, we develop a novel statistical method that can quantitatively measure the replicability of biomarkers by jointly analyzing prognostic and predictive effects. In simulation studies, our method shows improved power with well-controlled Fdr when compared with traditional meta-analysis methods (FE, RE) and MAMBA under different data generation processes. In the robustness analyses of our ground truth model, MAJAR is robust to different correlation relationships between the G and GT effects and varied magnitudes of outliers. We also observ that MAJAR performed similarly to MAMBA when only the G effect is present, which validates that MAJAR simplifies into MAMBA in that condition (i.e., without GT interaction effect). In terms of the computation time benchmarking, MAJAR has very efficient performance comparable to RE and MAMBA and converged within a minute under most simulation settings. In the real data application of the IMPROVE-IT PGx GWAS data, we used a five-fold cross-validation strategy for demonstrating meta-analysis using MAJAR across multiple studies, which is effective in evaluating the method we proposed (although not optimal). There may exist other alternatives to classify subgroups of subjects based on a specific single study. Overall speaking, we find that MAJAR not only demonstrates some level of consistency with results from MAMBA and the pooled GWAS, but also boosts the power of association test and identifies more SNPs with numerous biological evidence. In summary, MAJAR is an efficient tool that can provide statistical evidence of whether the discovered biomarkers are replicable without using an independent replication dataset. The identified replicable biomarkers may provide more insights for biological follow-up and drug development, for example, with more confident support for patient stratification, drug response prediction, or late-stage biomarker-guided adaptive trial designs.

One limitation of MAJAR is that we assume the latent statuses of the prognostic and predictive effects are the same, which means the identified replicable variants only represent the variants with joint effects. If only one effect exists in the real data, MAJAR simplifies into a marginal test like MAMBA, and can only detect marginal effects (Figure S4). Another limitation is that the ground truth model of MAJAR assumes independence across SNPs. For computational considerations, we recommend that users clump and prune the variants before inputting them into our workflow to follow the current model assumption of MAJAR and meanwhile boost the computational speed, especially under the GWAS setting with millions of variants. However, we didn’t explore the robustness of MAJAR if this assumption is violated in this study. It may be worthwhile exploring this in the future. Third, we used a unified *α* to characterize the outliers of all studies for both prognostic and predictive effects. This assumption may be violated if 1) there is outlier heterogeneity across different studies and 2) there are differences between the outliers from the prognostic effect and outliers from the predictive effect. Fourth, the current framework assumes the true effect size follows a normal distribution. This assumption may be violated if the real distribution of true effect size has a heavy tail. The performance of MAJAR under situations violating those assumptions needs further effort to explore.

Our proposed method can be extended in several directions. First, we can explore the performance of our method in broader settings. With the accumulating data from large biobanks such as UK biobank, the performance of our method for testing genotype and genotype-by-environment interaction on disease genetics is worth exploring. In addition, we only demonstrate the results in a setting with a large number of variants in this study, so application to settings with a small number of clinical or genomic biomarkers is another avenue to explore. Furthermore, our method can be extended to a two-dimensional space for testing prognostic and predictive effects, respectively. This could be realized by designing a framework with two latent variables for both prognostic and predictive effects (i.e., *R*_*G*_ and *R*_*GT*_, *O*_*G*_ and *O*_*GT*_), which enables us to do the interaction test conditional on the main effect.

In the era of precision medicine, many predictive biomarkers have been discovered for drug response prediction and patient stratification. However, validating these biomarkers has been challenging in many cases given the difficulty to find an independent replication dataset to validate the discovered signals. Our proposed MAJAR method provides an alternative approach by statistically calculating the posterior probability of replicability for identified biomarkers, which represents an important step in accelerating the usage of novel predictive biomarkers in clinical practice.

## Data Availability

Genotype data were processed using PLINK v2.00: https://www.cog-genomics.org/plink/1.9/; MAMBA R package: https://github.com/dan11mcguire/mamba; metafor R package v3.4.0: https://cran.r-project.org/package=metafor; Rcpp R package v1.0.9: https://cran.r-project.org/package=Rcpp; TMB R package v1.9.1: https://cran.r-project.org/package=TMB; figures were generated using ggplot2 R package v3.3.6: https://cran.r-project.org/package=ggplot2 and figures with break axis were generated using ggbreak package v0.1.1: https://cran.r-project.org/package=ggbreak; Manhattan plot was generated using qqman R package v0.1.8: https://cran.r-project.org/package=qqman; Venn diagram was generated using ggvenn R package v0.1.9: https://cran.r-project.org/package=ggvenn; upset plot was generated using UpSetR R package v1.4.0; GWAS Catalog: https://www.ebi.ac.uk/gwas; Open Targets: https://www.opentargets.org. Our method is implemented in the MAJAR R package, freely available at https://github.com/JustinaXie/MAJAR.

## Acknowledgments

We would like to thank Dr. Hong Zhang for helpful discussions and Dr. Rachel Marceau West and Dr. Lan Luo for their helpful comments on our manuscript.

## Key Points

- We proposed a novel method MAJAR (Meta-Analysis of Joint effect Associations for biomarker Replicability assessment) to jointly test prognostic and predictive effects and assess replicability of biomarkers without the need of using an independent replication study.
- Simulation results show MAJAR is robust to outliers, computationally efficient, and provides improved power with well-controlled Fdr compared with other methods.
- MAJAR can be applied to pharmacogenomics (PGx) GWAS, disease genetics studies, and other more general biomarker studies.

