## Supplemental Material for "Statistical Assessment of Biomarker Replicability using MAJAR Method"

**This file includes:**

Supplementary Methods

Figures S1 to S11

Table S1

References

**Other Supplementary Materials for this manuscript include the following:**

Table S2

**Supplementary Methods**

1. **Optimizing** $\boldsymbol{(}\boldsymbol{\tau}^{\boldsymbol{2}}\boldsymbol{,\rho)}$ **parameters**

We estimate parameters $\boldsymbol{\theta}=(p,\lambda,\alpha,\tau^{2},\rho)$ using an EM algorithm [1]. The derivation of complete data log-likelihood and E-step is provided by the supplementary information of MAMBA [2], so we only show the steps of optimizing $(\tau^{2},\rho)$ in details here.

To maximize the expected complete data log-likelihood on $\tau^{2}$ and $\rho$, we need to integrate $\boldsymbol{\mu}_{\boldsymbol{j}}=(\mu_{j,G},\mu_{j,GT})$

$$ll\left( \tau^{2},\rho\right)\propto Q\left( \tau^{2},\rho\right)=P\left( R_{j}=1 \right)\log(\int P\left( \boldsymbol{b}_{\boldsymbol{j}} | \boldsymbol{\mu}_{\boldsymbol{j}} \right)P\left( \boldsymbol{\mu}_{\boldsymbol{j}}|\tau^{2},\rho\right)d\boldsymbol{\mu}_{\boldsymbol{j}}).$$

(1)

By Bayes theorem,

$$P\left( \boldsymbol{b}_{\boldsymbol{j}} | \boldsymbol{\mu}_{\boldsymbol{j}} \right)P\left( \boldsymbol{\mu}_{\boldsymbol{j}}|\tau^{2},\rho\right)\propto P(\boldsymbol{\mu}_{\boldsymbol{j}}|\boldsymbol{b}_{\boldsymbol{j}},\tau^{2},\rho).$$

(2)

The posterior of $\boldsymbol{\mu}_{\boldsymbol{j}}\boldsymbol{|}\boldsymbol{b}_{\boldsymbol{j}}$ is $\mathrm{MVN}\left( m_{j},V_{j} \right),$

$$m_{j}\left( \tau^{2},\rho\right)=V_{j}\left( \sum_{k=1}^{K} \Sigma_{\mathrm{jk}}^{-1}b_{jk} \right),$$

$$V_{j}\left( \tau^{2},\rho\right)=\left( \sum_{k=1}^{K} \Sigma_{\mathrm{jk}}^{-1}+\Omega^{-1} \right)^{-1},$$

$$\Sigma_{jk}=\left[ \begin{matrix} s_{jk,G}^{2} & 0 \\ 0 & s_{jk,GT}^{2} \end{matrix} \right], \Omega=\tau^{2}\left[ \begin{matrix} 1 & \rho\\ \rho& 1 \end{matrix} \right].$$

With the fact that$\int P\left( \boldsymbol{\mu}_{\boldsymbol{j}} | \boldsymbol{b}_{\boldsymbol{j}},\tau^{2},\rho\right)d\boldsymbol{\mu}_{\boldsymbol{j}}\boldsymbol{=}1$, we can sort out different terms related to $\tau^{2}$ and $\rho$ in $P\left( \boldsymbol{b}_{\boldsymbol{j}} | \boldsymbol{\mu}_{\boldsymbol{j}} \right)P\left( \boldsymbol{\mu}_{\boldsymbol{j}}|\tau^{2},\rho\right)$ and $P(\boldsymbol{\mu}_{\boldsymbol{j}}|\boldsymbol{b}_{\boldsymbol{j}},\tau^{2},\rho)$ to solve the integration in (1).

Then, the expected log-likelihood in round $(t+1)$ can be approximated by

$$\sum_{j=1}^{M} \hat{R}_{j}^{\left( t \right)}(-\log\left| \hat{\Omega}^{\left( t \right)} \right|-\log\left| \left( \hat{V_{j}^{-1}} \right)^{\left( t \right)} \right|+{m_{j}^{\left( t \right)}}^{'}\left( V_{j}^{-1} \right)^{\left( t \right)}m_{j}^{\left( t \right)}),$$

(3)

where $V_{j}^{-1}=\sum_{j=1}^{K} \Sigma_{jk}^{-1}+\Omega^{-1}=\left[ \begin{matrix} \sum_{k=1}^{K} \frac{1}{s_{jk,G}^{2}}+\frac{1}{\tau^{2}(1-\rho^{2})} & -\frac{\rho}{\tau^{2}(1-\rho^{2})} \\ -\frac{\rho}{\tau^{2}(1-\rho^{2})} & \sum_{k=1}^{K} \frac{1}{s_{jk,GT}^{2}}+\frac{1}{\tau^{2}(1-\rho^{2})} \end{matrix} \right].$

2. **Simulation procedures for fixed effect model**
1) Simulate the latent variables $R_{j}$ and $O_{jk}$

$R_{j}\sim\mathrm{Bernoulli}\left( \pi\right), O_{jk}\sim\mathrm{Bernoulli}\left( \lambda\right)$.

2) Simulate the true effect sizes from a spike-and-slab distribution

$\left( \begin{matrix} \mu_{j,G} \\ \mu_{j,GT} \end{matrix} \right)|\left( R_{j} \right)=\left\{ \begin{matrix} MVN(0,\Omega) & R_{j}=1 \\ (0,0)' & R_{j}=0 \end{matrix} \right.$, where $\Omega=\left[ \begin{matrix} \tau^{2} & \rho\tau^{2} \\ \rho\tau^{2} & \tau^{2} \end{matrix} \right]$.

3) Simulate $b_{jk}$ given $\mu_{j}$

$\left( \begin{matrix} b_{jk,G} \\ b_{jk,GT} \end{matrix} \right)|\left( \begin{matrix} \mu_{j,G} \\ \mu_{j,GT} \end{matrix} \right)\sim MVN(\left[ \begin{matrix} \mu_{j,G} \\ \mu_{j,GT} \end{matrix} \right],\Sigma=\left[ \begin{matrix} s_{jk,G}^{2} & 0 \\ 0 & s_{jk,GT}^{2} \end{matrix} \right])$.

3. **Simulation procedures for random effect model**

1) and 2) same as MAJAR and FE

3) Simulate the $\eta_{jk,G} \mathrm{and} \eta_{jk,GT}$ from the DerSimonian and Laird method [2,3] test statistics

$X_{G}=\frac{\left( K-1 \right)\sum_{j} \frac{1}{s_{jk,G}^{2}}}{(\sum_{j} {\frac{1}{s_{jk,G}^{2}})}^{2}-\sum_{j} \left( \frac{1}{s_{jk,G}^{2}} \right)^{2}}$, $X_{GT}=\frac{\left( K-1 \right)\sum_{j} \frac{1}{s_{jk,GT}^{2}}}{(\sum_{j} {\frac{1}{s_{jk,G}^{2}})}^{2}-\sum_{j} \left( \frac{1}{s_{jk,GT}^{2}} \right)^{2}}$,

$$\omega_{G}^{2}=\max\left( 0,\frac{I^{2}}{1-I^{2}}*X_{G} \right),\omega_{GT}^{2}=\max\left( 0,\frac{I^{2}}{1-I^{2}}*X_{GT} \right),$$

$$\eta_{jk,G}\sim N\left( \mu_{j,G},\omega_{G}^{2} \right),\eta_{jk,GT}\sim N\left( \mu_{j,GT},\omega_{GT}^{2} \right),$$

where

- $\eta_{jk,G}$ and $\eta_{jk,GT}$are the study-specific prognostic and predictive effect in cohort k for variant j,
- $\mu_{j}$ is the overall population mean genetic effect,
- $I^{2}$ indicates heterogeneity among studies,
- $\omega_{G}^{2}$ and $\omega_{GT}^{2}$ are the variance of study-specific prognostic and predictive effects, characterizing heterogeneity across study cohorts.

4) Simulate $b_{jk}$ given $\eta_{jk, G} \mathrm{and} \eta_{jk,GT}$

$\left( \begin{matrix} b_{jk,G} \\ b_{jk,GT} \end{matrix} \right)|\left( \begin{matrix} \eta_{jk,G} \\ \eta_{jk,GT} \end{matrix} \right)\sim MVN(\left[ \begin{matrix} \eta_{jk,G} \\ \eta_{jk,GT} \end{matrix} \right],\Sigma=\left[ \begin{matrix} s_{jk,G}^{2} & 0 \\ 0 & s_{jk,GT}^{2} \end{matrix} \right])$.

**Supplementary Figures**

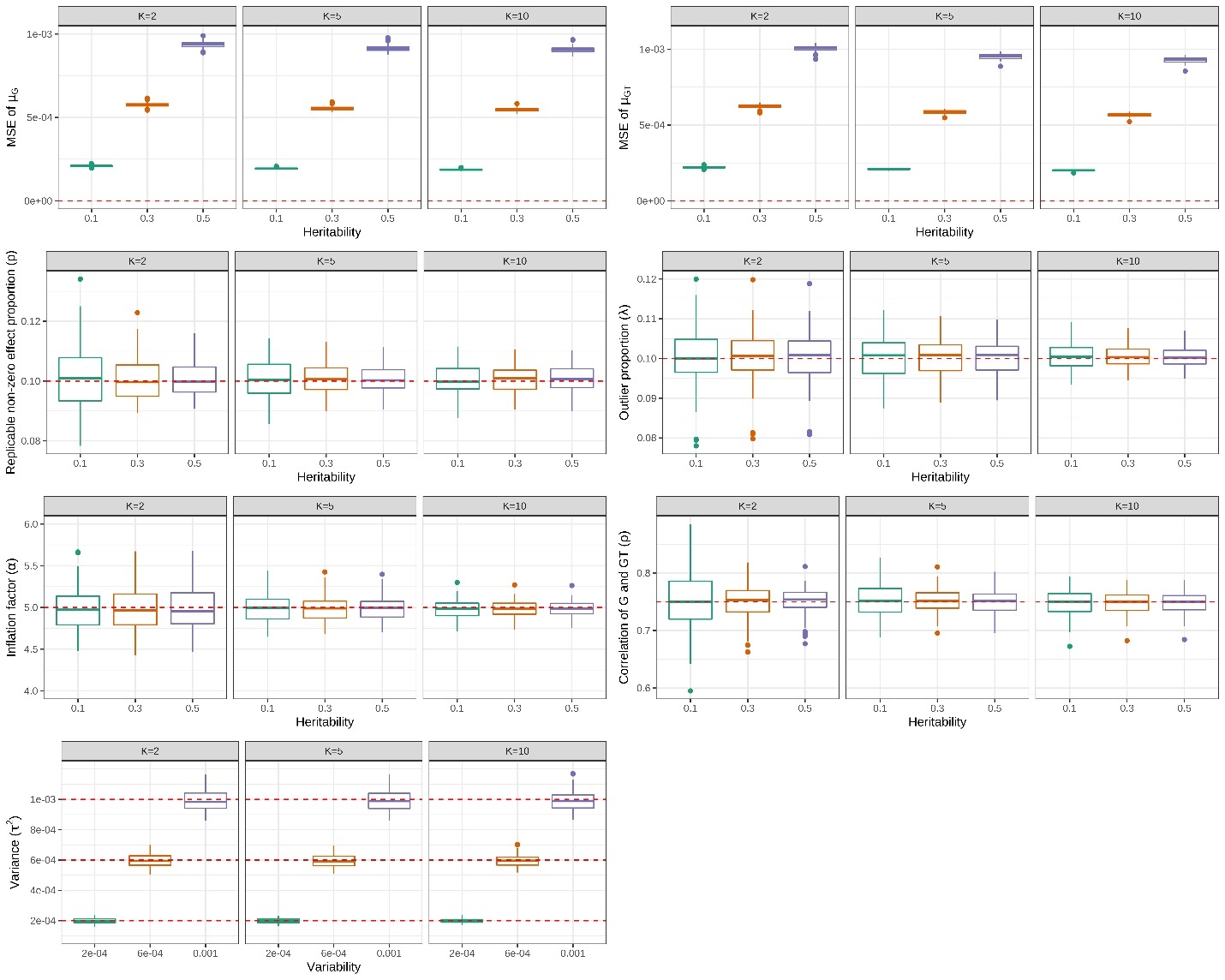

**Figure S1. MAJAR estimated parameters well under the MAJAR ground truth model.** For each panel, the columns represent the number of studies $K$, and the x-axis represents the heritability/variability ($\tau^{2}=h^{2}/Mp$) of the G and GT effects. The first two panels in the top row show the MSE of $\mu_{G}$ and $\mu_{GT}$, respectively. With the increase in heritability, the variance of the G and GT effects $\tau^{2}$ also increases, so the MSE of $\mu_{G}$ and $\mu_{GT}$ increases. The next five panels in the middle and bottom rows show the estimation performance of the proportion of replicable non-zero effect SNPs $p$, outlier proportion $\lambda$, inflation factor $\alpha$, correlation between G and GT effects $\rho$ and variance $\tau^{2}$ with the increase of heritability/variability. The estimated values of these parameters in 100 replications under each simulation setting are visualized by the colored boxplots. The red dashed lines represent the true values of these parameters under the ground truth model.

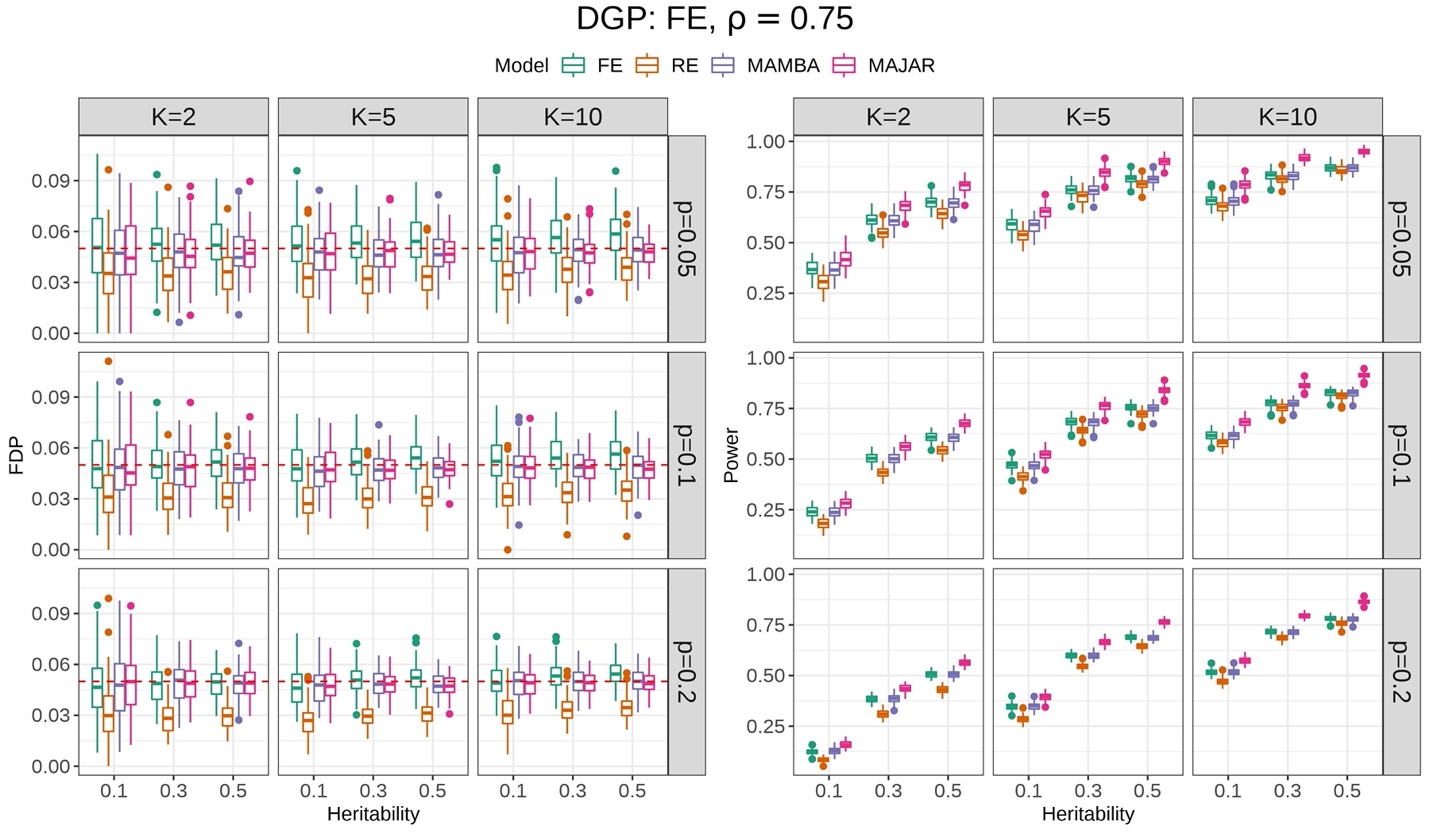

**Figure S2. Simulation result comparisons for FE, RE, MAMBA, and MAJAR under the FE model.** The data generation process is FE and $\rho$ is set at 0.75 in this simulation. The left panel is for the FDP performance, and the right panel is for the power performance. For each panel, the columns represent the number of studies $K$, and the rows represent the proportion of replicable non-zero effect SNPs $p$. Each panel shows the performance of FDP and power (y-axis) with increasing heritability $h^{2}$ (x-axis) under different settings of $K$ and $p$. The performance of each method in 100 replications under each simulation setting is visualized by the colored boxplots. The red dashed lines in the FDP panel represent the target Fdr threshold of 0.05.

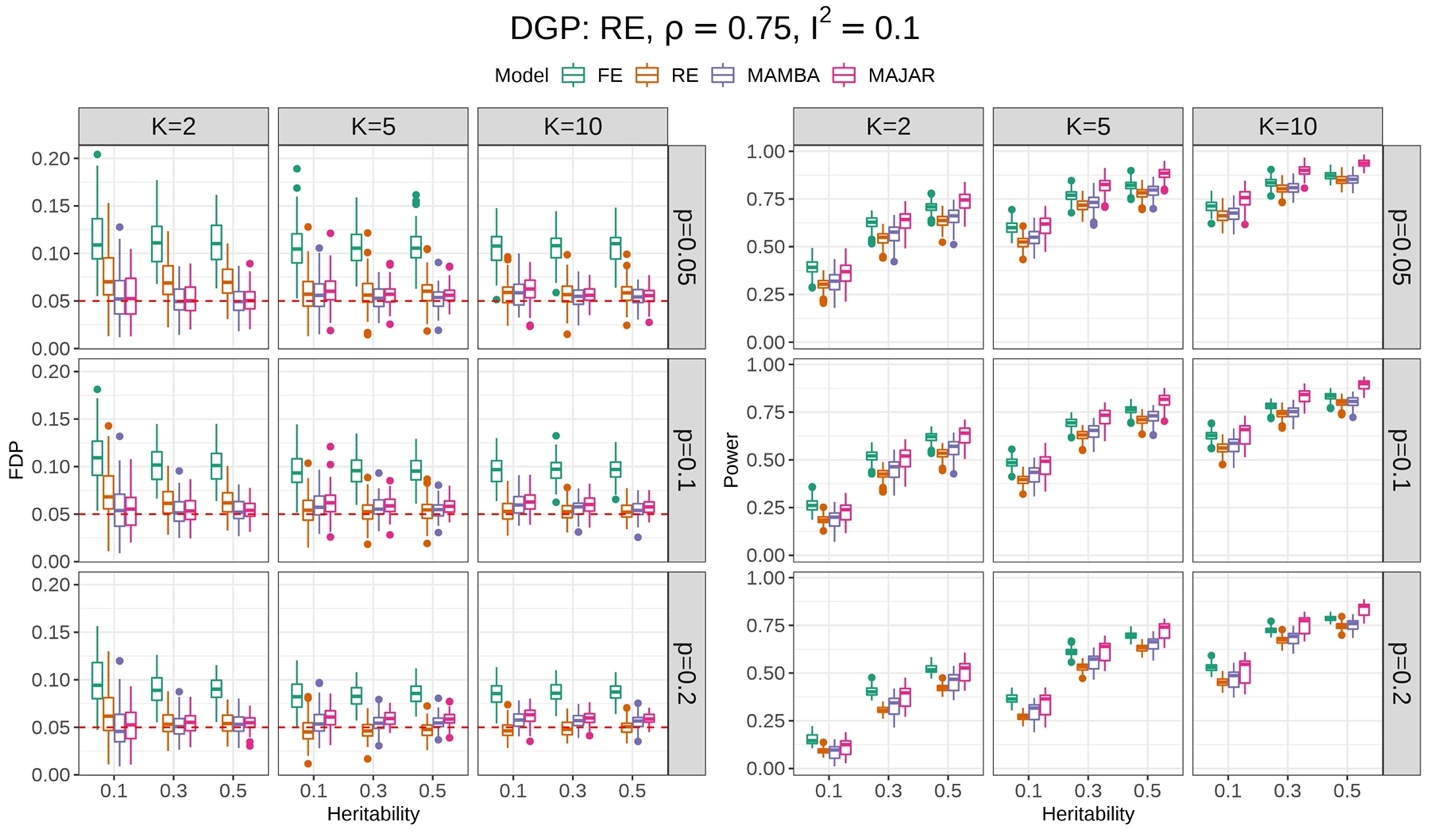

**Figure S3. Simulation result comparisons for FE, RE, MAMBA, and MAJAR under the RE model.** The data generation process is RE, $\rho$ is set at 0.75 and the $I^{2}$ is set as 0.1 in this simulation. The left panel is for the FDP performance, and the right panel is for the power performance. For each panel, the columns represent the number of studies $K$, and the rows represent the proportion of replicable non-zero effect SNPs $p$. Each panel shows the performance of FDP and power (y-axis) with increasing heritability $h^{2}$ (x-axis) under different settings of $K$ and $p$. The performance of each method in 100 replications under each simulation setting is visualized by the colored boxplots. The red dashed lines in the FDP panel represent the target Fdr threshold of 0.05.

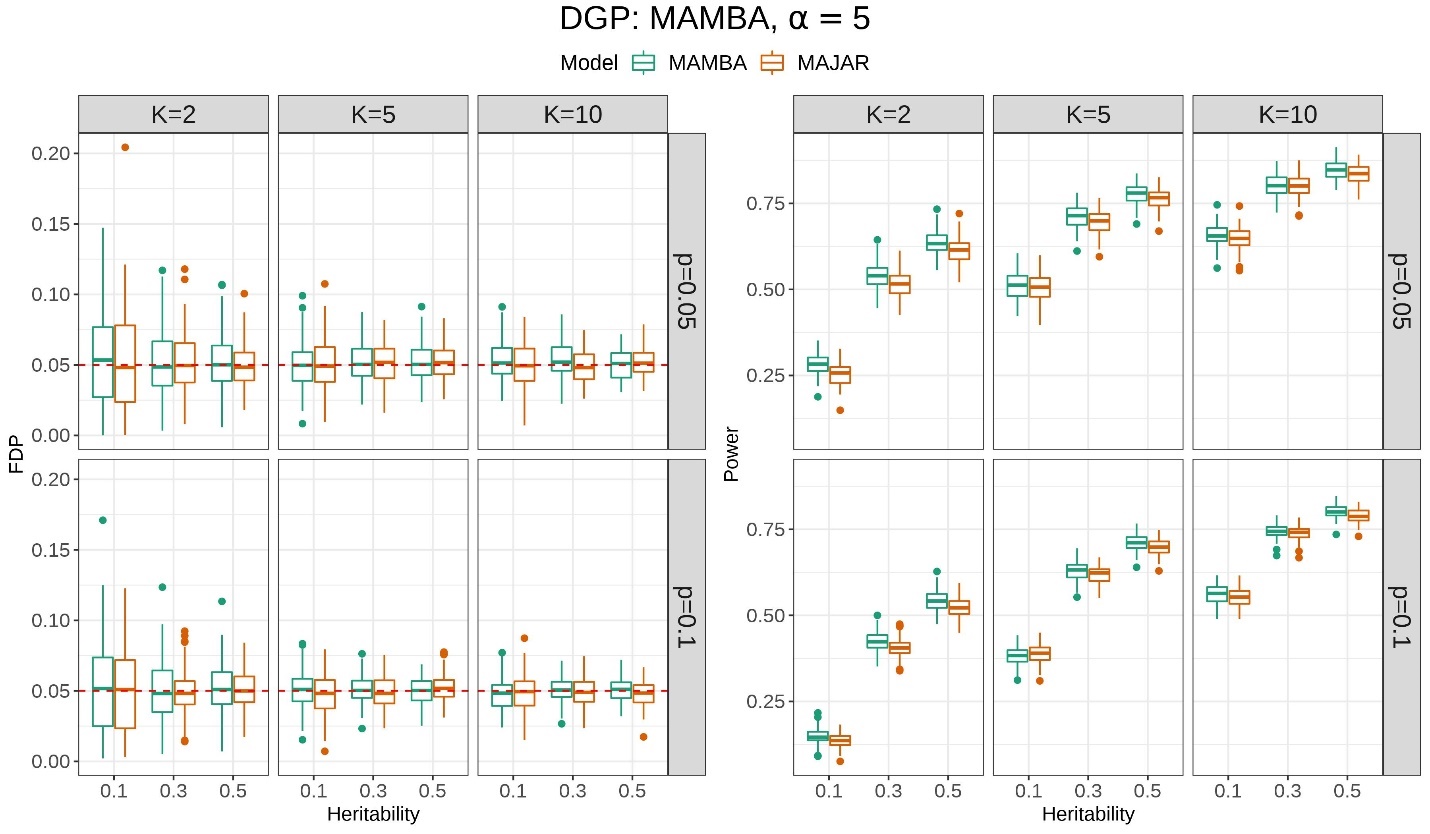

**Figure S4.** **Simulation results of FDP and power while comparing MAMBA and MAJAR when only the G effect presents.** The data generation process is MAMBA and $\alpha$ is set at 5. The left panel is for the FDP performance, and the right panel is for the power performance. For each panel, the columns represent the number of studies $K$, and the rows represent the proportion of replicable non-zero effect SNPs $p$. Each panel shows the performance of FDP and power (y-axis) with increasing heritability $h^{2}$ (x-axis) under different settings of $K$ and $p$. The performance of MAMBA and MAJAR in 100 replications under each simulation setting is visualized by the colored boxplots. The red dashed lines in the FDP panel represent the Fdr threshold of 0.05.

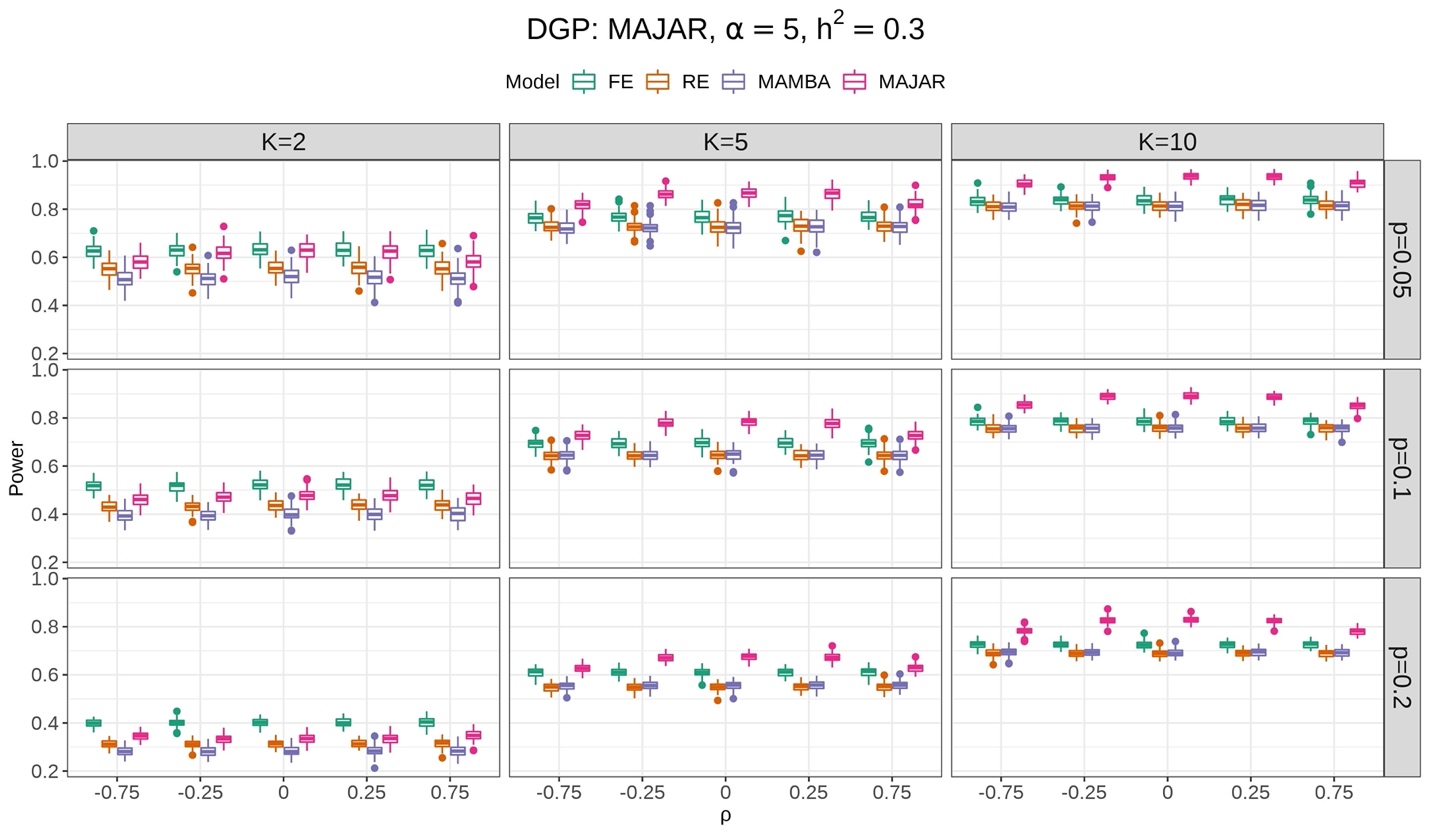

**Figure S5. MAJAR is robust to different correlation relationship between G and GT effects.** The data generation process is MAJAR, $\alpha$ is set at 5 and the heritability $h^{2}$is set at 0.3 in this simulation. The columns represent the number of studies $K$, and the rows represent the proportion of replicable non-zero effect SNPs $p$. Each panel shows the change of power (y-axis) with increasing correlation between G and GT effects $\rho$ (x-axis) under different settings of $K$ and $p$. The power performance of FE, RE, MAMBA, and MAJAR in 100 replications under each simulation setting is visualized by the boxplots.

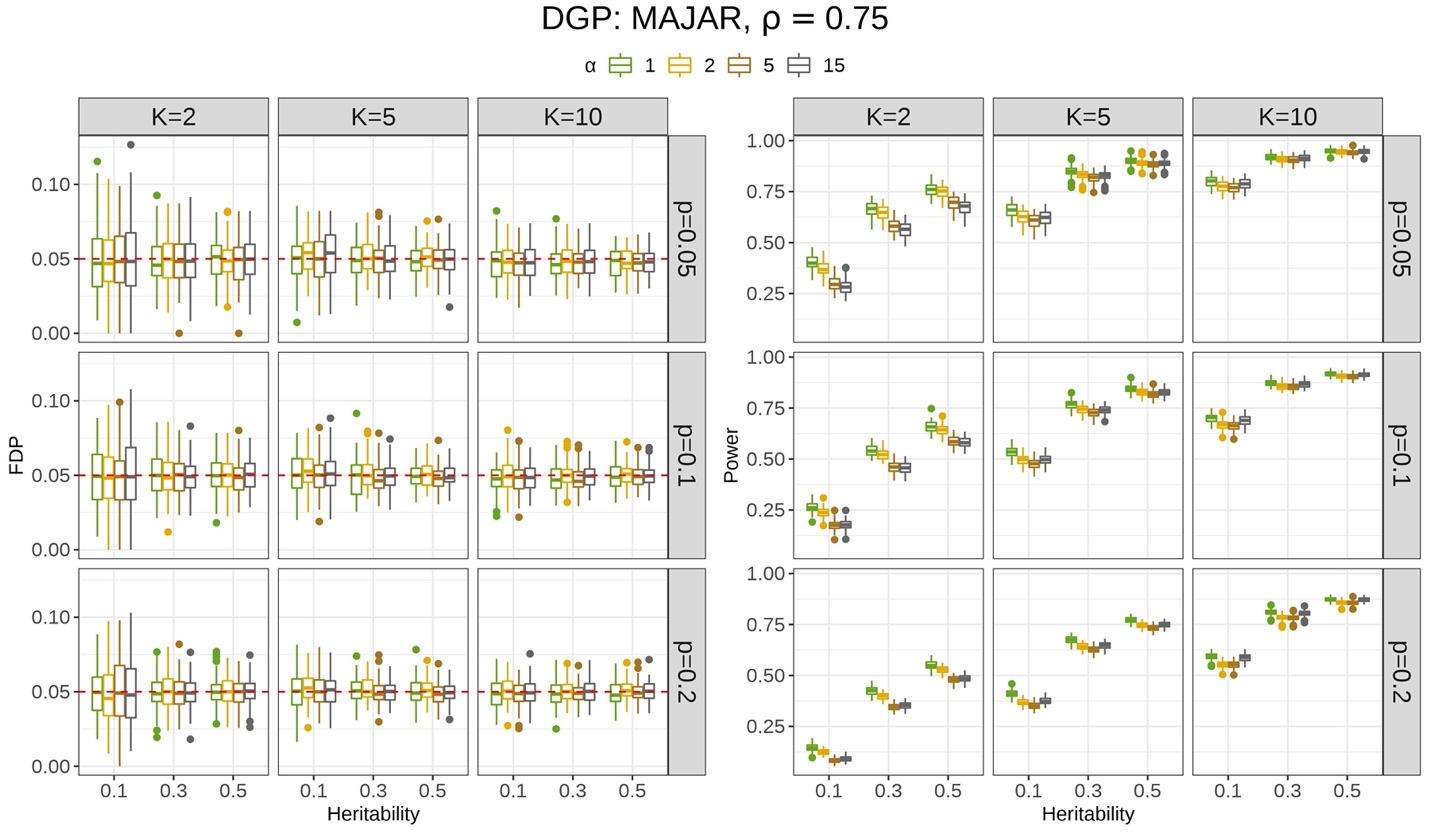

**Figure S6. MAJAR is robust to different levels of outliers.** The data generation process is MAJAR and ρ is set at 0.75 in this simulation. The left panel is for the FDP performance, and the right panel is for the power performance. For each panel, the columns represent the number of studies $K$, and the rows represent the proportion of replicable non-zero effect SNPs $p$. Each panel shows the performance of FDP and power (y-axis) with increasing heritability $h^{2}$ (x-axis) under different settings of $K$ and $p$. The performance of MAJAR under different levels of inflation factor $\alpha$ in 100 replications under each simulation setting is visualized by the colored boxplots. The red dashed lines in the FDP panel represent the target Fdr threshold of 0.05.

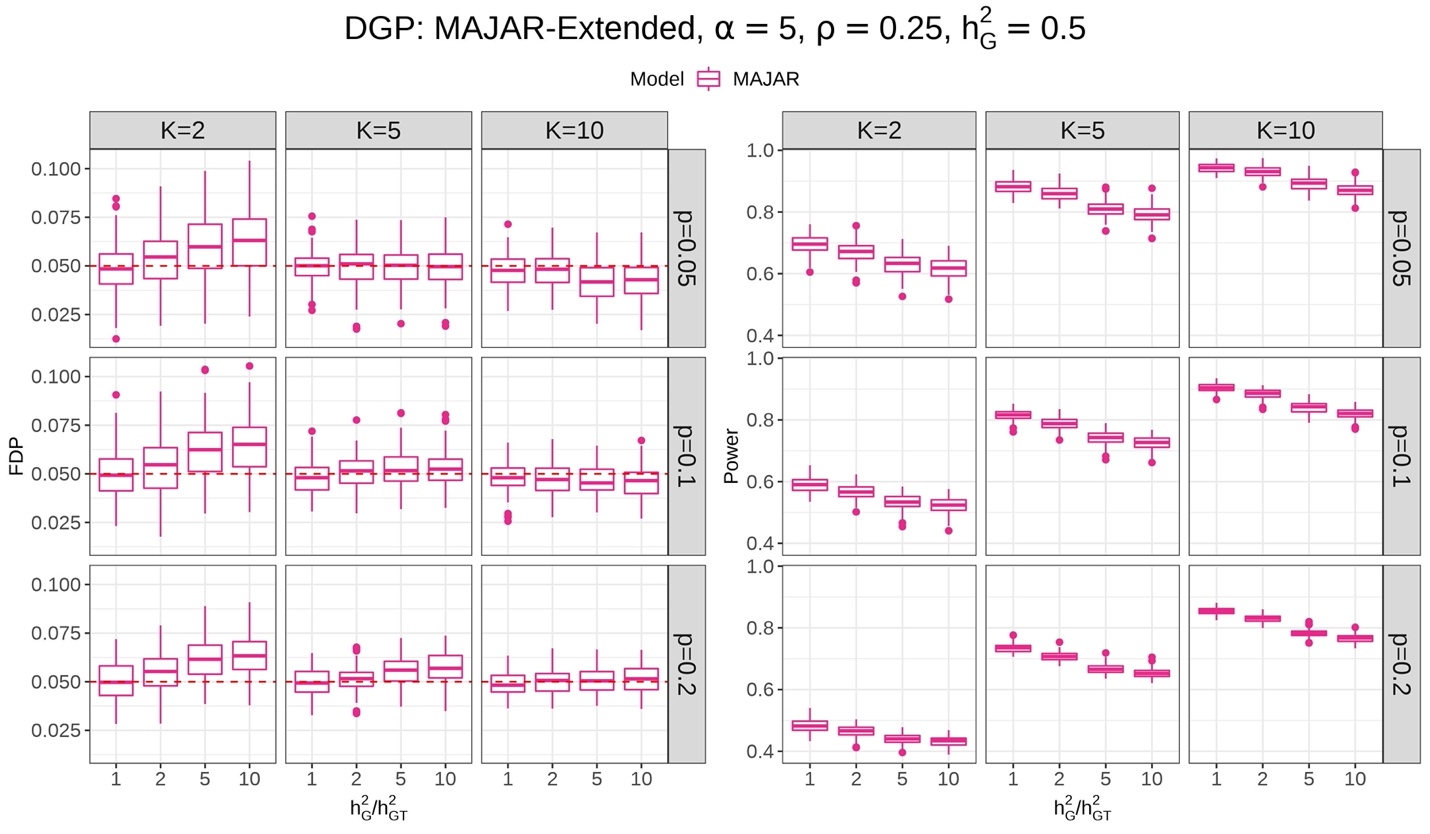

**Figure S7. MAJAR is robust to different scales of variance between G and GT effects (i.e., specified in the extended model).** The data generation process is MAJAR extended model, $\alpha$ is set at 5, $\rho$ is set at 0.25, and the heritability $h_{G}^{2}$is set at 0.5 in this simulation. The left panel is for the FDP performance, and the right panel is for the power performance. For each panel, the columns represent the number of studies $K$, and the rows represent the proportion of replicable non-zero effect SNPs $p$. Each panel shows the performance of FDP and power (y-axis) with increasing the ratio of heritability $\frac{h_{G}^{2}}{h_{GT}^{2}}$ (x-axis) under different settings of $K$ and $p$. The performance of MAJAR in 100 replications under each simulation setting is visualized by the boxplots. The red dashed lines in the FDP panel represent the target Fdr threshold of 0.05.

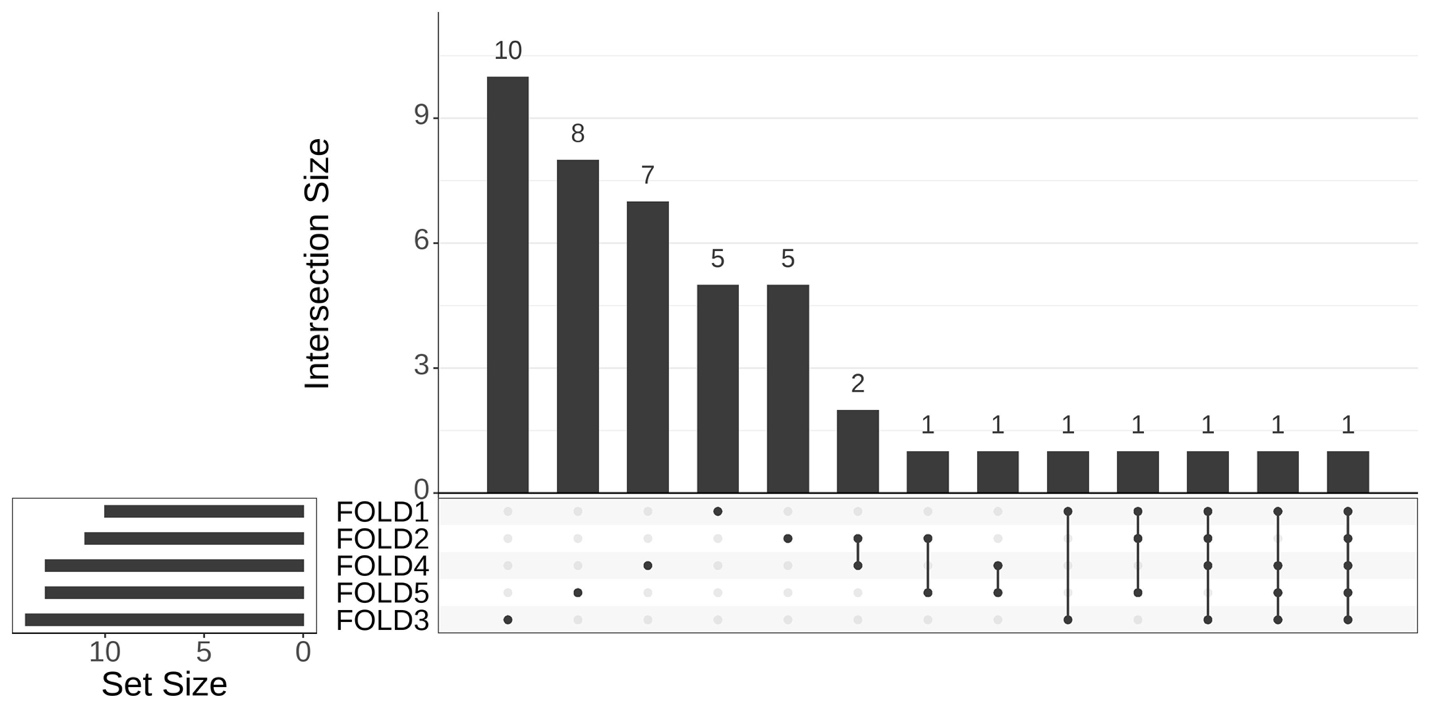

Chr17: rs56746789

Chr19: rs1065853

Chr9: rs62531463

**Figure S8. Upset plot for the 5-fold cross-validation robustness analysis of the IMPROVE-IT PGx GWAS data.** The vertical red line represents the boundary of variants that are identified in at least 4 leave-one-out analyses. The top three variants with PPR > 0.99 (rs1065853, rs62531463, rs56746789) in the 5-fold analysis are robust in the leave-one-out analyses.

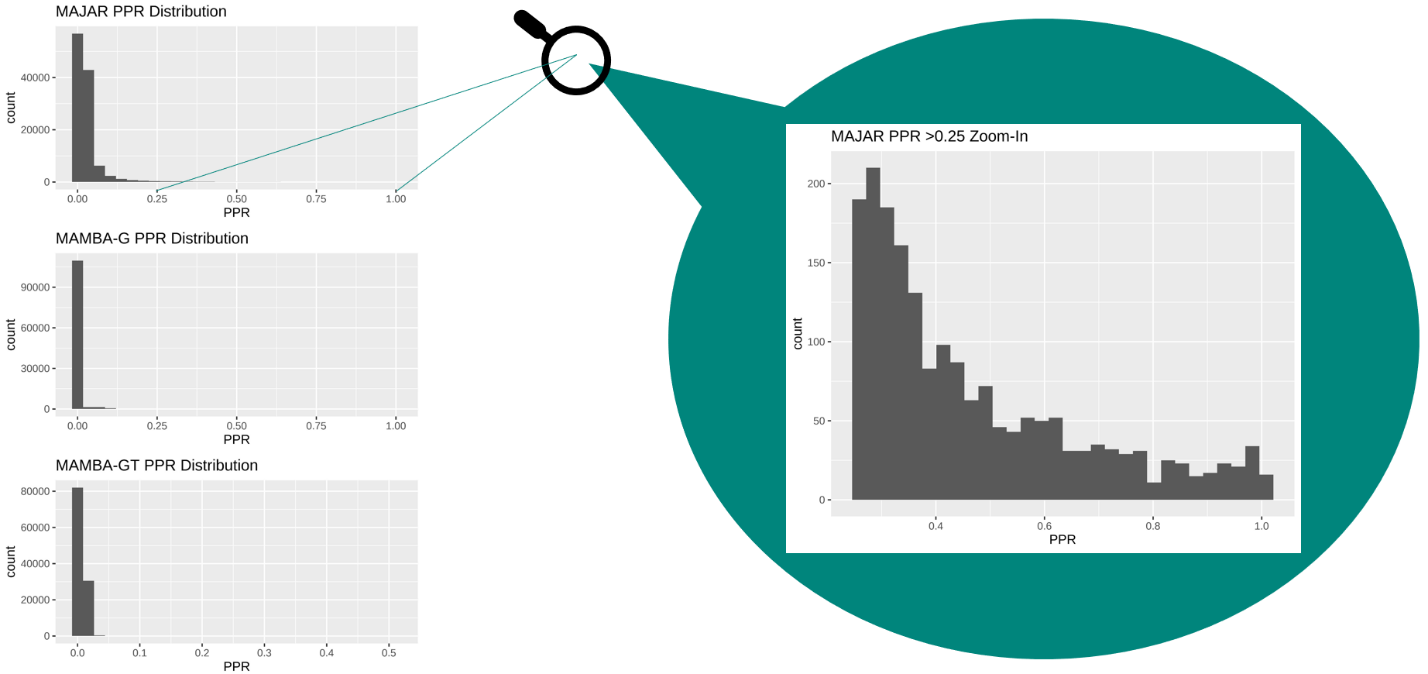

**Figure S9. Histograms of MAJAR PPRs, MAMBA-G PPRs, and MAMBA-GT PPRs for SNPs from the IMPROVE-IT PGx GWAS data analysis across the whole genome (after LD clumping and pruning).** All three PPRs show a right-skewed distribution with long right tail, indicating that most SNPs are not replicable (non-zero effect) signals. The corresponding frequency drops more smoothly when PPR increases for MAJAR method, compared to MAMBA-G and MAMBA-GT.

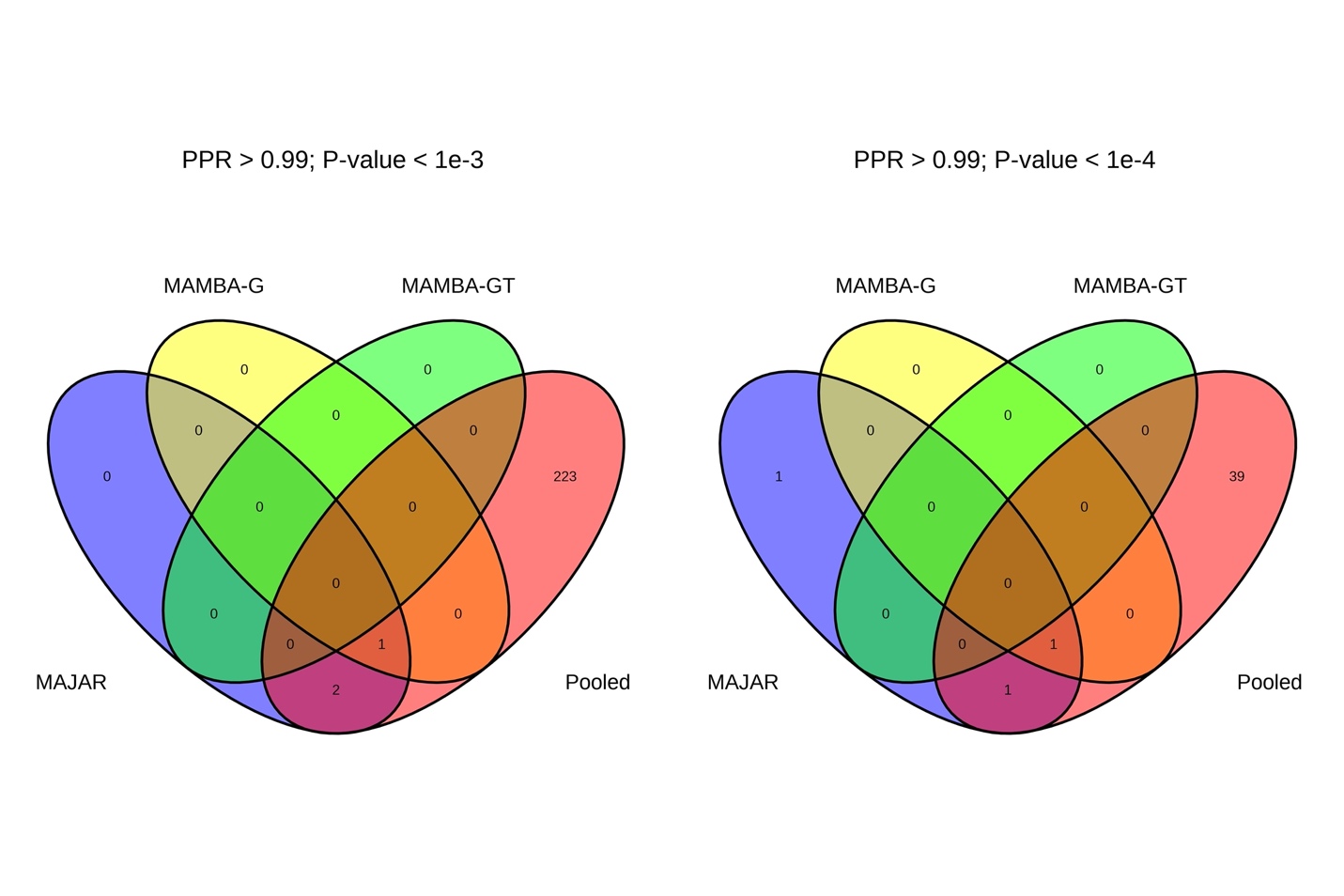

**Figure S10. Venn diagrams of variants identified by MAJAR, MAMBA-G, MAMBA-GT, and 2-df test from the pooled PGx GWAS summary statistics in the analysis of IMPROVE-IT PGx GWAS data.** The left panel illustrates the numbers of the overlapping and different variants across MAJAR PPR > 0.99, MAMBA-G PPR > 0.99, MAMBA-GT PPR > 0.99, and 2df test p-values of the pooled GWAS <$1\times{10}^{-3}$. The right panel illustrates the numbers of the overlapping variants across MAJAR PPR > 0.99, MAMBA-G PPR > 0.99, MAMBA-GT PPR > 0.99, and 2df test p-values of the pooled GWAS < $1\times{10}^{-4}$.

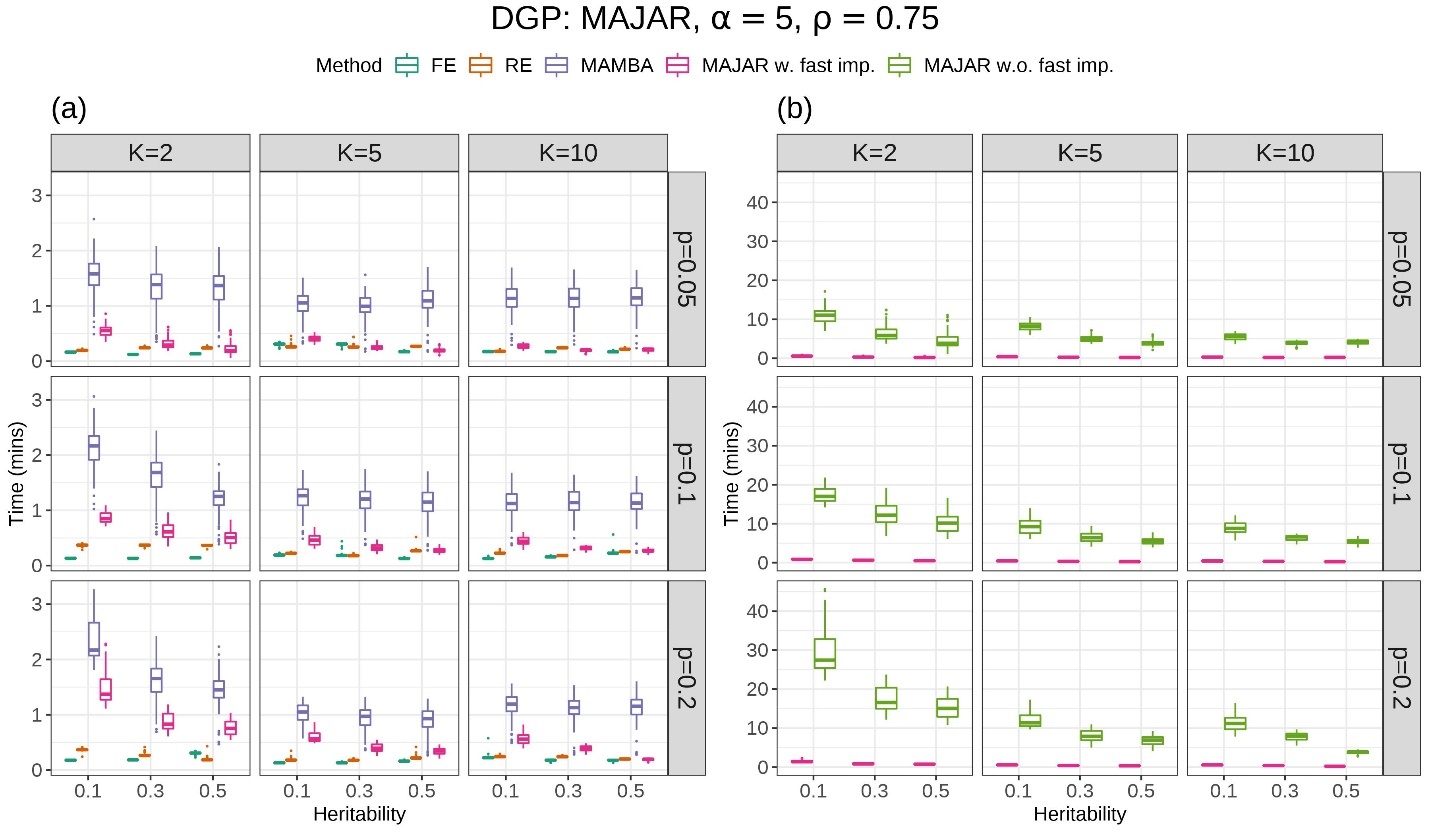

**Figure S11. Computation time comparison.** (a) Among FE, RE, MAMBA, MAJAR with fast implementation (MAJAR w. fast imp.). (b) between MAJAR with fast implementation and MAJAR without fast implementation (MAJAR w.o. fast imp.). The data generation process is MAJAR, $\alpha$ is set at 5 and the $\rho$ is set as 0.75 in this simulation. The columns represent the number of studies $K$, and the rows represent the proportion of replicable non-zero effect SNPs $p$. Each panel shows the performance of computation time in minutes (y-axis) with increasing heritability $h^{2}$ (x-axis) under different settings of $K$ and $p$. The time performance of each method in 100 replications under each simulation setting is visualized by the colored boxplots.

**Table S1. Top variants identified by MAJAR with Fdr < 0.05 in the analysis of IMPROVE-IT PGx GWAS data and their results and annotation information.**

| **CHR** | **POS** | **A1** | **A2** | **RSID** | **Gene Symbol/**  **Nearest Coding Gene** | **MAJAR PPR** | **MAJAR Fdr** | **MAMBA-G PPR** | **MAMBA-GT PPR** | **CCT**  ***p*-values** | **Pooled GWAS  2df *p*-values** | **Pooled GWAS  GT *p*-values** | **Pooled GWAS  G *p*-values** |
| --- | --- | --- | --- | --- | --- | --- | --- | --- | --- | --- | --- | --- | --- |
| 19 | 45413233 | G | T | rs1065853 | *APOE/APOC1* | 1.00 | 1.38E-12 | 1.00 | 0.36 | 5.12E-14 | 1.09E-51 | 1.43E-4 | 3.79E-16 |
| 17 | 66629197 | CT | C | rs56746789 | *LINC01482* | 1.00 | 3.96E-04 | 0.83 | 0.51 | 1.05E-02 | 1.71E-05 | 7.79E-6 | 3.45E-05 |
| 9 | 517446 | T | G | rs62531463 | *KANK1* | 1.00 | 4.35E-03 | 0.64 | 0.44 | 5.17E-05 | 4.20E-04 | 1.20E-4 | 1.19E-03 |
| 21 | 28730882 | T | A | rs76451912 | *LOC124905003* | 0.99 | 1.35E-02 |  | 0.21 | 4.28E-05 | 3.40E-04 | 1.05E-4 | 7.62E-04 |
| 2 | 135165813 | T | G | rs4954144 | *MGAT5* | 0.98 | 1.56E-02 | 0.52 | 0.21 | 1.81E-02 | 1.63E-04 | 5.96E-5 | 4.00E-04 |
| 21 | 16487700 | A | G | rs2823068 | *LOC107985483* | 0.98 | 1.90E-02 | 0.73 | 0.06 | 7.14E-02 | 7.20E-04 | 1.84E-4 | 1.94E-03 |
| 2 | 235349598 | C | T | - | *-* | 0.97 | 2.21E-02 | 0.67 | 0.05 | 6.66E-02 | 5.06E-04 | 1.35E-4 | 1.15E-03 |
| 4 | 163970150 | T | C | rs1351220 | *NAF1* | 0.97 | 3.25E-02 | 0.49 | 0.28 | 5.46E-02 | 6.67E-04 | 2.19E-4 | 1.04E-03 |
| 2 | 202475180 | CA | C | rs35421171 | *C2CD6* | 0.97 | 2.63E-02 | 0.43 | 0.21 | 3.41E-03 | 7.88E-04 | 1.70E-4 | 3.22E-03 |
| 8 | 123477319 | T | C | rs35903937 | *SMILR* | 0.96 | 3.68E-02 | 0.64 | 0.03 | 6.53E-02 | 3.52E-04 | 2.49E-3 | 6.97E-05 |
| 15 | 23038344 | G | A | rs28510330 | *NIPA2* | 0.96 | 4.29E-02 | 0.37 | 0.10 | 3.46E-02 | 6.78E-04 | 4.79E-4 | 3.94E-04 |
| 15 | 89399986 | C | G | rs201822759 | *ACAN* | 0.96 | 4.34E-02 | 0.37 | 0.11 | 2.17E-02 | 1.10E-03 | 2.78E-4 | 2.36E-03 |
| 9 | 118728024 | T | G | rs61381472 | *PAPPA* | 0.95 | 2.91E-02 | 0.51 | 0.08 | 7.21E-03 | 7.66E-04 | 1.55E-4 | 5.53E-03 |

**Table S2. (Separate file) Evidence of Mapped Genes of 13 Identified Variants from GWAS Catalog.**
